# Excess mortality in Germany during 2020–2023: A descriptive age-stratified analysis

**DOI:** 10.64898/2026.06.16.26355759

**Authors:** Martin Sauter

## Abstract

This study investigates excess mortality in Germany in the years from 2020 to 2023 and its temporal alignment with reported COVID-19 deaths. The analysis uses annual and weekly all-cause mortality data and linear baseline trends derived from pre-pandemic years. Possible effects of demographic and population changes on baseline trends were also examined. Excess mortality was analysed over time and across age groups.

Excess mortality was observed in all investigated years, rising from 2020 to its highest value in 2022. In absolute terms, the age group ≥80 years accounted for the largest proportion of excess deaths throughout the study period. After 2021, elevated mortality relative to baseline was also observed in younger age groups down to 15 years of age, although absolute numbers remained substantially lower than in older groups. No evidence of excess mortality was observed for individuals younger than 15 years.

Periods of excess mortality were temporally aligned with waves of reported COVID-19 deaths. In 2020, cumulative excess mortality after calendar week 11 closely matched reported COVID-19 deaths (43 876 vs. 41 835 deaths). Weekly excess mortality, reported COVID-19 deaths and wastewater viral load, when available showed strong temporal synchrony, although excess mortality increasingly exceeded reported COVID-19 deaths during later pandemic waves. Temporal patterns differed from the typical seasonal mortality peaks commonly associated with influenza epidemics during the early months of the year. In 2023, excess mortality declined substantially, possibly indicating a return to mortality levels before the emergence of SARS-CoV-2.

## 1 Introduction

### 1.1 General

The first infection with the SARS-CoV-2 virus in Germany was detected in January 2020 in Munich [1]. The increased spread of infections occurred from mid-March, presumably due to vacationers returning from the ski regions of Austria [2], leading to rapid rise of infections and subsequent the first contact restriction measures at the end of March 2020.

The first outbreaks of the COVID-19 pandemic in Northern Italy, Wuhan, and New York had already shown that primarily older people faced a significantly increased risk of death from infection with SARS-CoV-2 [3], [4], [5]. The initial outbreak in Germany in late March 2020 did not lead to the catastrophic conditions and the number of deaths seen in New York or Bergamo with more than even 10-fold increases of death numbers during the same time [6], [7], but only in some moderate excess mortality as we will see later.

There is no doubt that the recording of deaths in its entirety is highly reliable and precise, providing one of the best possible foundations for quantifying the effects of the COVID-19 pandemic. These aspects are of both public and scientific interest.

#### 1.2.1 Introduction and Definition

Excess mortality is defined as the degree to which the actual number of deaths exceeds the expected number of deaths. In common demographic calculations, there are different numbers which may be used for an assessment:

1. Crude Death Count (CDC) refers to the absolute number of deaths in a population over a specific time period.
2. Crude Mortality Rate (CMR) is a measure of the number of deaths per people in a population
3. Age-Standardized Mortality Rate (ASMR) is a mortality rate that has been adjusted to account for differences in the age distribution of the population. Age dependent CMRs are applied to a defined standard population [8].

#### 1.2.2 Calculation of Excess Mortality

Many works, especially the ones comparing many countries use CDC, as these values are publicly available by national statistic offices and calculation is straight forward [9], [10], [11], [12]. A smaller number of authors use CMR or ASMR [13], [14], [15], which require more data work.

A counterfactual approach is typically based on extrapolation from mortality trends observed in previous years. In many studies, this baseline is estimated using linear extrapolation from pre-pandemic years. Depending on the quantity which is extrapolated, different trends have to be considered: In most of the developed high-income countries, population is aging and CDC is rising due to demographic shift.

This approach is used in the “World Mortality Dataset” by Kobak and Karlinsky[12], which is also implemented on the “Our World in Data / Excess Mortality”[10] project website using the pre-pandemic years from 2015 to 2019.

The “Human Mortality Database” [16] offers a similar interactive online tool, albeit with many more options for calculating weekly excess mortality. Users can select reference years or choose adjust the type of reference (median, trend, average etc.)

Various national statistic offices did not extrapolate by rising trends, but instead median values of previous years or averages which leads to underestimation of the baseline and overestimation of deaths [17], [18].

When ASMR is used [15] falling trends can be seen and have to be extrapolated, here linear decrease is also used as approximation.

There were also attempts by international institutes and organisations, namely WHO and IHME, which tried to assess the global death toll of the pandemic [11], [9]. These authors used splines for extrapolation. However, curvature on the edges may lead to extrapolation exceeding or strongly damping the linear trend and were questioned if they were appropriate as models [19].

EuroMoMo uses Poisson regression for modelling of annual trends instead of linear regression [20]. Because annual mortality in Germany has a very linear characteristic exhibiting only minimal curvature during the pre-pandemic period, the exponential growth assumed by Poisson regression closely approximates a linear trend. Consequently, the choice between linear and Poisson regression has only a negligible effect on the resulting excess mortality estimates. This is discussed in section S1 of the Supplementary Material as well.

Other studies estimate expected deaths using actuarial approaches that combine mortality rates from life tables with population numbers from national statistical offices [21] [22] [23]. The resulting baseline estimates also show approximately linear trends, although with higher predicted mortality levels than the baseline used here. A comparison of these approaches with the present analysis is also provided in the Supplementary Material section S1.

More advanced methods try to calculate values of expected deaths during the year and take seasonal effects into account. Seasonal effects are modelled by empirical approaches using splines with different parameters [20] or a decomposition of seasonal trends into Fourier series [13]. Anyway, more complex calculations require more underlying assumptions regarding regular phases of excess mortality events (Influenza waves in winter and heat waves in summer). For these cases, the annual trends are often modelled by a linear approach, the seasonal dependencies often by splines.

In many commonly used excess mortality frameworks annual mortality trends are nevertheless extrapolated using linear approaches, whereas seasonal variation is modelled separately. In the present study, a linear baseline model using the years 2013–2019 was therefore chosen because pre-pandemic mortality in Germany showed a stable approximately linear increase during this period while requiring comparatively few modelling assumptions. The detailed analysis in the supplement shows that the trend derived from 2013 to 2019 is the median of the seven possible trends going back from 2019.

## 2 Applied Methods

### 2.1 Materials and Calculation Tools

This study uses publicly available data. Death numbers from Germany for various age bands, sex and ages for the relevant years are collected and further distributed by the German National Statistics Office (NSO) Destatis, offering an interactive database for annual deaths by sex [24] and age [25]. Population Numbers were also taken from Destatis. [26]. During the pandemic “special issues” (“Sonderauswertung”) about current mortality was published weekly [17], but in a rather machine-unfriendly format as an excel spreadsheet. Other sites like Eurostat also redistribute these data [27], [28], same for the “human mortality database” [29]. Format and aggregation in form of the age bands vary and it is comfortable to choose the data format most comfortable for least coding effort.

The relevant source for certified COVID-19 deaths is German Public-Health Institute Robert Koch-Institut (RKI). Data on COVID-19 fatalities was distributed in different formats as Excel-spreads until mid of 2023. These datasets are not publicly available anymore and had been deleted from RKI’s web site, but are still accessible via wayback machine [30], [31].

The newer version is redistributed via github [32], and continues until 2024. Destatis also offers annual death causes reports, which also includes information about fatalities by COVID-19 [33]. The problem is that all of these data sources do not match with each other and give slightly different values. An overview is given in the supplementary materials. For the annual and weekly data from 2020 to 2022, the source from RKI [31] was used. Annual data was aggregated from monthly values. For weekly data in 2023, the later version[34] was used. For annual data in 2023, the values of RKI [34] were listed in Table 1 for best consistency. There are differences, but these do not affect the overall outcome of this study. A detailed listing of these values can be found in supplemental materials, S 5.

**Table 1.**
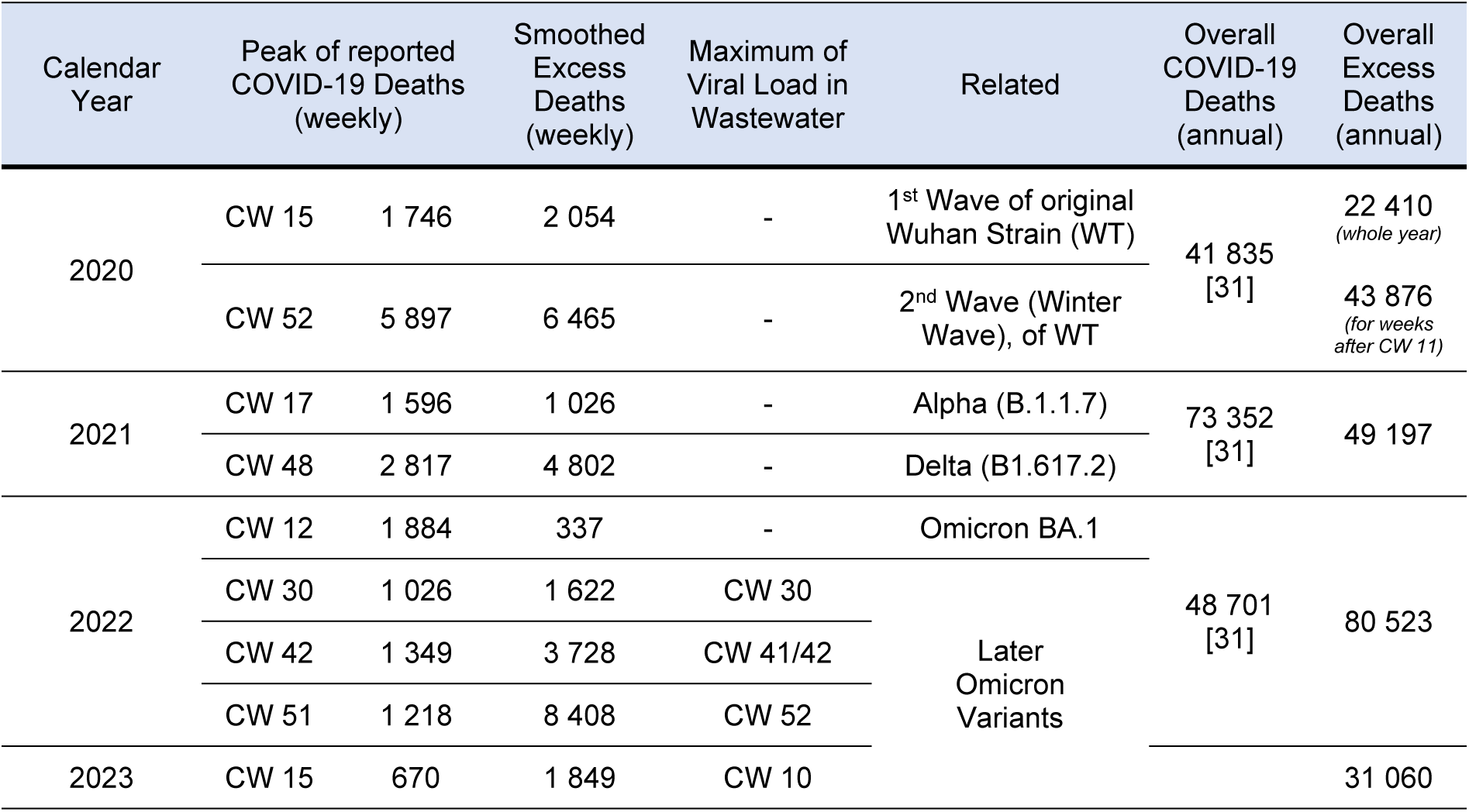
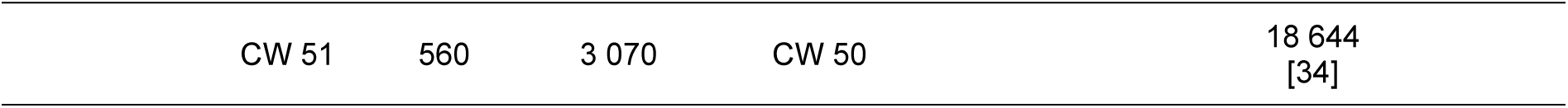
Extracted quantitive key parameters for the years 2020-2023. Central reference is the peak of reported COVID-19 deaths (column 3, with the relevant calendar week mentioned in column 2). Corresponding excess deaths in the same week are shown in column 4, wastewater signal after May 2022 in column 5). Column 6 gives the attribution to the reported wave and the relevant variants (when known).Column 7 and 8 summarize COVID-19 and excess deaths for the whole year; as a special case for 2020 excess deaths after time of first larger virus intrusion (CW 11) are also summarized.

Age dependent data of COVID-19 fatalities come from RKI, and are best listed in this reference [30]. RKI also publishes wastewater surveillance data (“Amelag” Project) via github for download [35].

Plotting and calculations where done using Python3 [36], NumPy [37], Matplotlib[38], the Python Scipy [39] and scikit-learn linear model [40] packages.

### 2.2 Basic calculations

Median value for prediction comes from the linear trend from the years 2013-2019, this fit which will serve as baseline for all other analyses. The reason for this trend is that it is gives the median values of prediction when evaluating the possible trends going backwards from 2019 to 2010 to 2016. Detailed values can be found in Supplements Section S 1.

The analytical expression for this trend is:

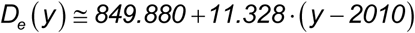

Basic strategy about calculation of deviation and error estimation comes here by the relative deviation between expected *D_E_(y)* and observed deaths *D_O_(y)* in the year, which is usually called “P-score” [41]:

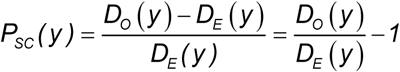

In Section S 6 from the supplemental materials an age-standardised calculation of relative excess is done. Outcome strongly depends as well on the age bands and the standard population chosen – 5-year age-bands with the standard population of Germany in 2020 matches the outcomes in terms of the P-score quite well, but using wider bands or younger populations shift relative results from away from the CDC model used here.

### 2.3 Error estimation

Maximum P-score from the previous years is used to decide whether CDC is above expectation bounds or within it. Maximum and minimum values are also shown in Figure 1. In 2014 there is a relative deviation of –3% towards the trend whereas 2015 shows the maximum deviation towards higher values of +2.06%. All of the discussed years are above the 2.06%, with 2022 and 2021 showing the highest values.

**Figure 1:**
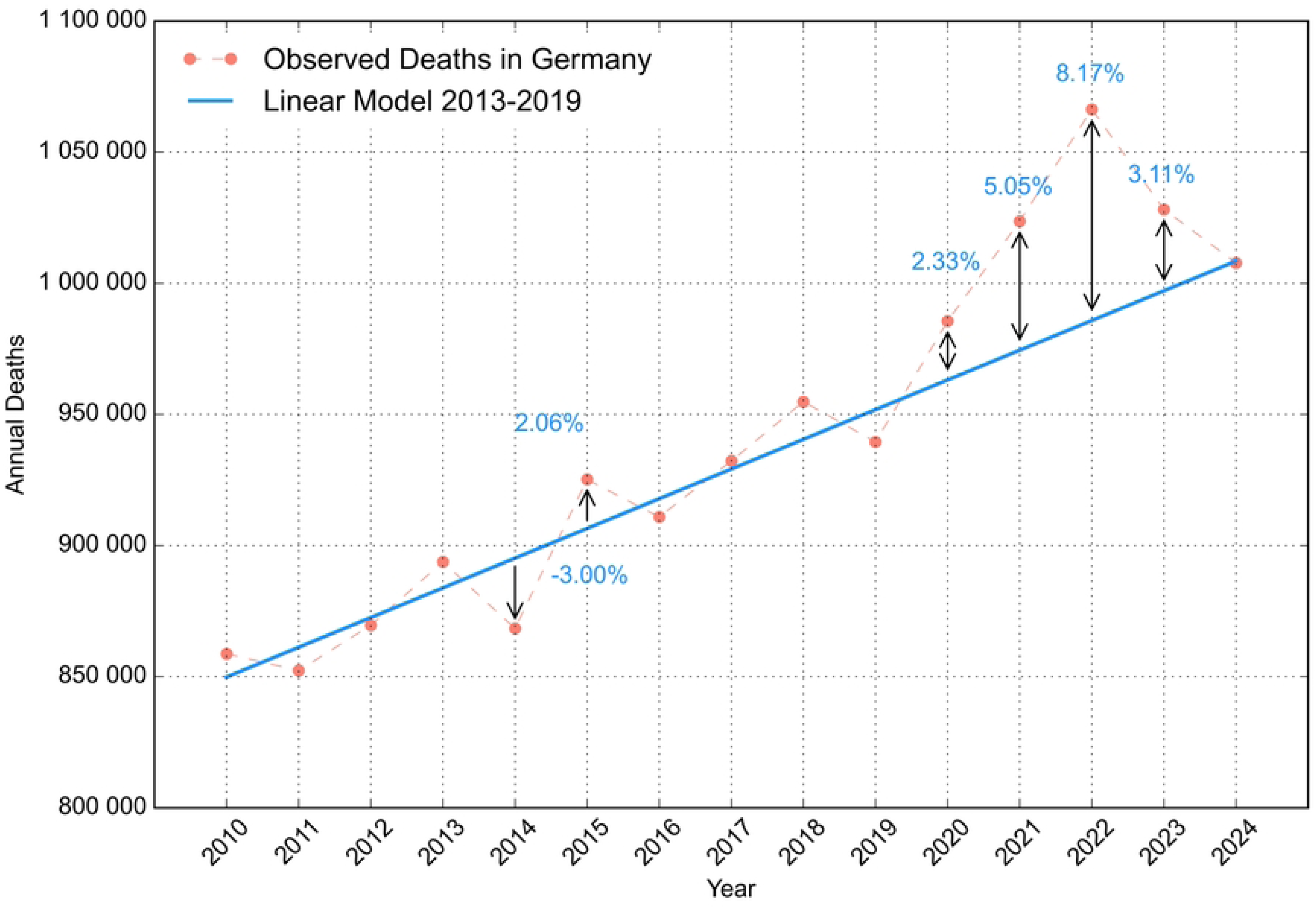
Observed Deaths from 2010 until 2024, drawn together with the fit of the linear regression resulting from timespan 2013-2019. Relative minimum and maximum deviations in the reference periods are shown, as well as for all years after 2019.

How these values match with PIs calculated for linear models is shown in S 2. The deviations calculated here are not symmetrical, unlike PIs, which assume a normal distribution of the data underlying.

### 2.4 Decomposition of the linear regression into subgroups

Linear regression applies to a set of a dependent variables *y_i_*and independent variables *x_i_* (*i=1…N*). Slope *m_y_*and intercept *b_y_* are calculated for least squares and predict values from extrapolation.

Now, the set *(x_i_, y_i_)* is split into two subsets *(x_i_, u_i_) and (x_i_, v_i_)* with *y_i=_ u_i+_ v_i_* while keeping *x_i_*the same. When calculating the linear regression parameters *(m_u_*, *b_u_)* and *(m_v_*, *b_v_)* for each of these two subsets it can be shown that:

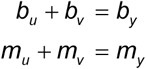

In consequence, the sum of extrapolations from each of the subgroups will predict the sum of the extrapolation from the full group. In reversal, splitting a set into subgroups can split up the predictions while keeping the sum. A short mathematical proof comes in chapter S 3 of the supplements.

If this applicable on a partition into two groups, it also works for further partitions. I will use this for division into weekly time intervals in section 3.1 and age groups in section 3.2.

## 3 Results and Discussion

### 3.1 Time-based assessments for the different years

Results for 2020 and the following years are shown in Figure 2. Each year is visualised by two panels, the upper shows reported deaths in red and the calculated baseline (2013-2019 trend) is given in blue dashed lines. The projected minimum/maximum deviation calculated from the previous years (calculated in the same way as in Section 2.3) and is shown as grey shaded area.

**Figure 2.**
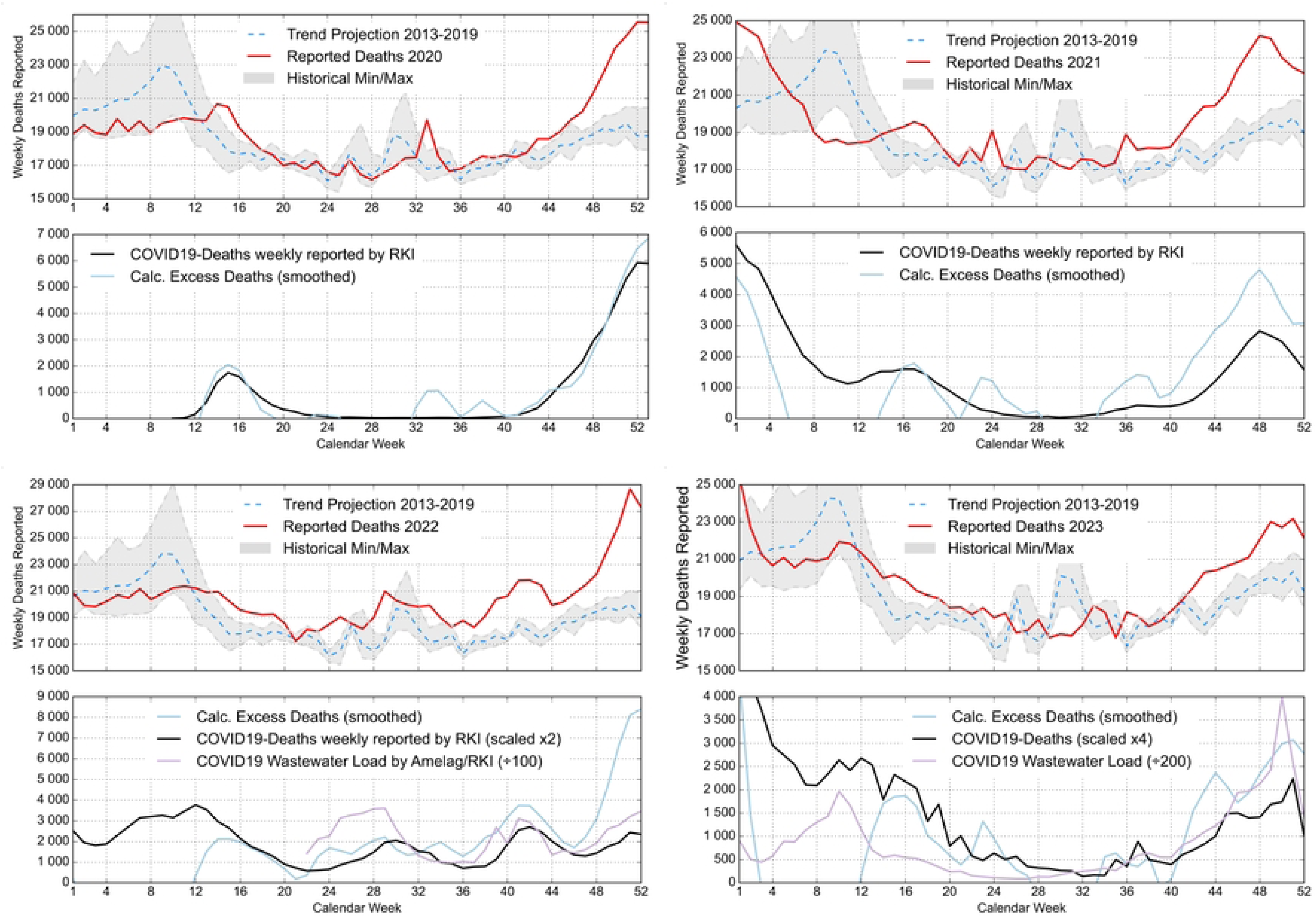
Reported and calculated excess mortality in weekly resolution for the four years discussed here. For each year, the upper panel shows reported deaths as red line, calculated expectation from the trend of 2013-2019 is shown as dashed blue line. Upper and lower deviations from the trends are shown as grey dashed lines, the area between the two is shaded grey. The lower panel shows the calculated and smoothed excess deaths in light blue, the reported deaths due to COVID-19 are shown in black (the y-axis in the lower panel is always truncated to zero at the lower boundary for better clarity. For the later years of 2022 and 2023, the reported COVID-19 deaths are scaled by factors of 2 or 4 resp. to better show close temporal alignment. For 2022 and 2023 viral wastewater load tracked by surveillance measurements has been added to the lower panel. The corresponding Sars-CoV2 variants associated with the waves are summarized in Table 1.

The lower panels show excess deaths, calculated as the difference between reported deaths and the estimated baseline, displayed as a light blue line. Excess mortality curves were smoothed using a Savitzky–Golay filter with a window length of 7 points and a third-order polynomial. Reported COVID-19 deaths from the German public health institute RKI (Section 2.1) are shown as a black line. For the years 2022 and 2023, COVID-19 death counts were scaled by factors of 2 and 4, respectively, to improve visualization of the temporal alignment between both curves.

Since May 2022, the AMELAG system established by RKI measured viral load in wastewater all over Germany [35]. These curves were added to the lower panels (b) as light violet line; a scaling of the viral load (measured in Copies/l) by a factor of 200 was also done to visualize time-dependent matching between wastewater load and the other two quantities.

The time-dependent effects are summarized in Table 1.

All different waves of the virus and its later subtypes can be identified. Each wave of COVID-19 deaths coincided with a wave of excess deaths. For 2020, the lines match very well. Almost all of the found waves (except the first wave of Omicron BA.1 subtype in 2022) do not happen in quarter 1 of the years, which is the season for the regular annual Influenza waves. This shows that SARS-CoV-2 has a different seasonality than Influenza and also the other common human coronaviruses, which come usually in the first quarter of the year [42].

For later pandemic waves, excess deaths exceeded reported COVID-19 deaths. This was particularly apparent during the Delta wave in autumn 2021, when the peak of excess deaths was almost twice as high as the corresponding number of reported COVID-19 deaths.

Later, the ratio between the annual values and the weekly peaks go down, shown in Figure 2 and in Table 1.

The year of 2022 is also of special interest as this year comes with the highest number of excess deaths. There are four waves of COVID-19 deaths and excess deaths; the first wave does not come with a clear excess mortality peak because the maximum coincides with a death peak resulting from Influenza waves in the former years. All of the following waves can be identified by a peak in viral wastewater load. Between these waves, deaths come higher than the calculated baseline for the whole year.

For 2023 there comes another shift in the pattern of excess and COVID-19 deaths, still showing a strong temporal coupling between these two. The first wave in 2023 comes again around early spring with a slightly shifted peak in excess mortality, similar to the first Omicron subtype wave in 2022. The second wave shows a rise of reported deaths from the baseline after CW 40, showing a rather long autumn wave. The wastewater signal increases in the same time. This reveals another transition in the pattern of COVID waves and attributed excess mortality.

Pearson cross-correlations were calculated between detrended and smoothed excess mortality and reported COVID-19 deaths in order to assess their temporal relationship for all four years investigated. Correlations were evaluated for time lags between −10 and +10 weeks using Pearson correlation coefficients. The lag with the maximum correlation coefficient was identified as the strongest temporal alignment between both time series. The calculated coefficients and time lags for best correlation are listed in Table 2.

**Table 2.** Calculated correlation coefficients and time lag for best correlations for the relationship between Excess Deaths and COVID-19 Deaths (Columns 2 and 3) and between Wastewater Viral Load and COVID-19 Deaths (Columns 3 and 4). For 2020 and 2021 there is no data for viral load available.

| Calendar<br>Year | Correlation between Excess Deaths<br>(smoothed) and COVID-19 Deaths |  | Correlation between COVID-19 Deaths and<br>Wastewater Viral Load |  |
| --- | --- | --- | --- | --- |
| | Max. Pearson $r$ | Lag at max. $r$ | Max. Pearson $r$ | Lag at max. $r$ |
| 2020 | 0.919 | 1 | - | - |
| 2021 | 0.962 | 0 | - | - |
| 2022 | 0.546 | -2 | 0.812 | -2 |
| 2023 | 0.693 | -1 | 0.808 | -1 |

The temporal correlation is very strong for 2020 and 2021 (max. r > 0.91). The correlation becomes moderate for 2022 and 2023 (r = 0.546 and 0.693, respectively).

For 2022 and 2023 the same calculation was done for viral load in wastewater and reported COVID-19 deaths. The calculated results are also shown in Table 2.

A lag of approximately two weeks between wastewater viral load and mortality is medically plausible and consistent with the expected delay between infection dynamics and COVID-19-related deaths. The correlation is moderately strong (r ∼ 0.81 for both years).

### 3.2 Excess Mortality in Age Groups

For this purpose, the population is split into age bands. Due to the database of the Destatis 6 age bands were chosen with the main intent to assure a continuous trend in population: 0-15, 25-29, 30-39, 40-59, 60-79 years and older than 80 years. The corresponding population development in the age groups is shown in Supplement section S 7. Trend of CDC is shown and the absolute numbers of excess deaths in the age groups is calculated.

#### 3.2.1 Absolute Number of Excess Deaths

The linear regression of CDC in the age groups is shown in Figure 3, resulting numbers are shown in Table 3.

**Figure 3.**
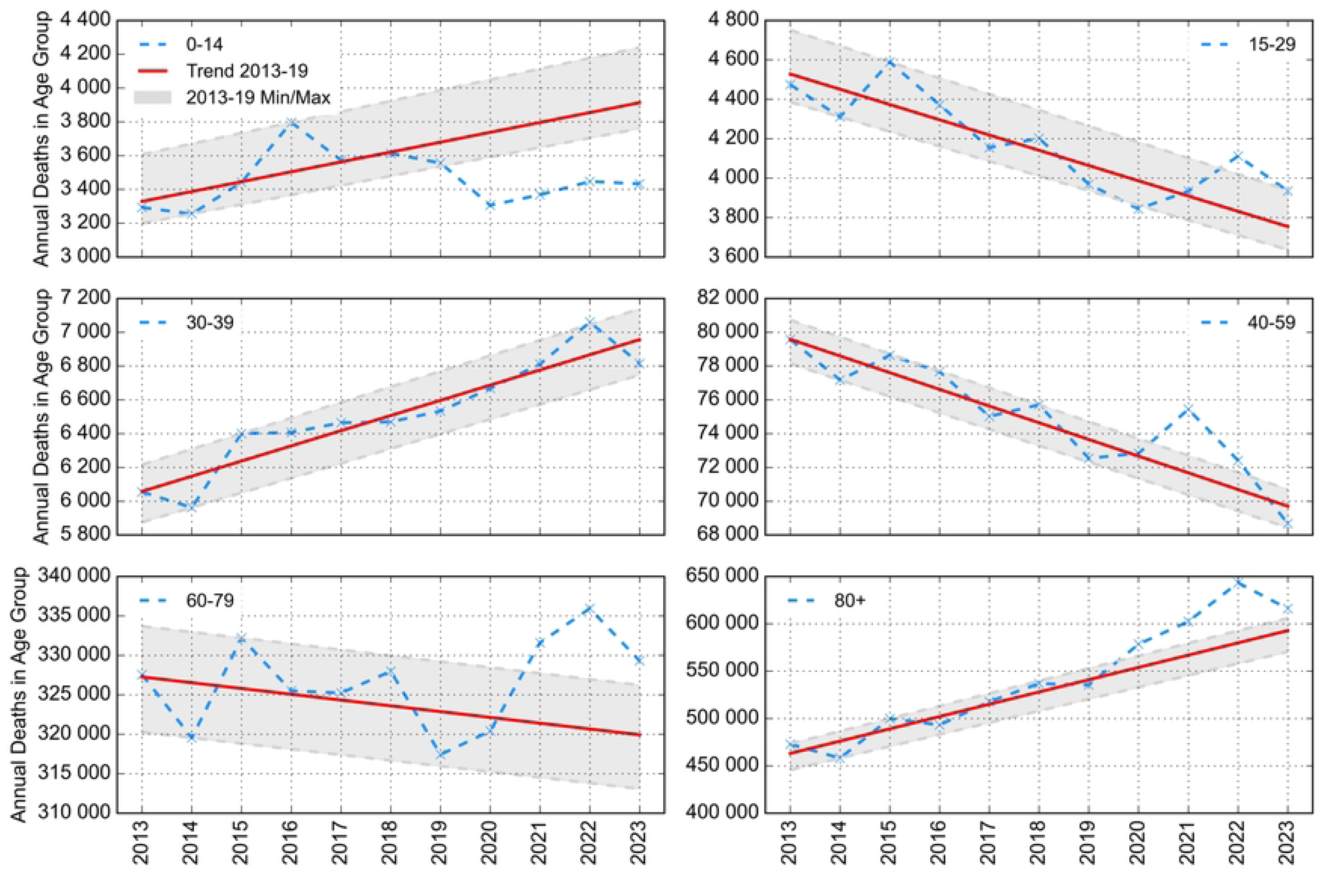
Deaths in reported age groups chosen here, used for excess death calculation. The baseline always comes from the linear trend from 2013 to 2019. Absolute and relative excess numbers calculated are summarized in Table 3.

**Table 3.**
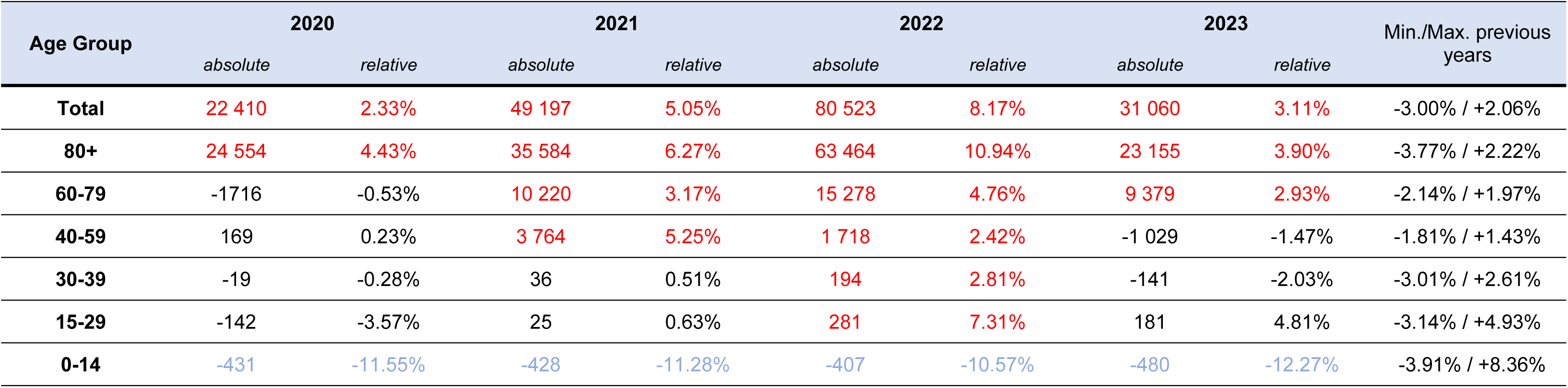
Absolute and relative numbers of excess deaths derived from the trend 2013-2019, here shown for the age groups used. Red numbers indicate excess mortality higher than the maximum of the previous years. When the reported deaths are lower than the relative minimum of the previous years (which is only happening for the group of age 0-14), numbers are in light blue.

| Age Group | 2020 |  | 2021 |  | 2022 |  | 2023 |  | Min./Max. previous years |
| --- | --- | --- | --- | --- | --- | --- | --- | --- | --- |
|  | absolute | relative | absolute | relative | absolute | relative | absolute | relative |  |
| Total | 22 410 | 2.33% | 49 197 | 5.05% | 80 523 | 8.17% | 31 060 | 3.11% | -3.00% / +2.06% |
| 80+ | 24 554 | 4.43% | 35 584 | 6.27% | 63 464 | 10.94% | 23 155 | 3.90% | -3.77% / +2.22% |
| 60-79 | -1716 | -0.53% | 10 220 | 3.17% | 15 278 | 4.76% | 9 379 | 2.93% | -2.14% / +1.97% |
| 40-59 | 169 | 0.23% | 3 764 | 5.25% | 1 718 | 2.42% | -1 029 | -1.47% | -1.81% / +1.43% |
| 30-39 | -19 | -0.28% | 36 | 0.51% | 194 | 2.81% | -141 | -2.03% | -3.01% / +2.61% |
| 15-29 | -142 | -3.57% | 25 | 0.63% | 281 | 7.31% | 181 | 4.81% | -3.14% / +4.93% |
| 0-14 | -431 | -11.55% | -428 | -11.28% | -407 | -10.57% | -480 | -12.27% | -3.91% / +8.36% |

The vast majority of excess deaths occurred in individuals older than 80 years throughout the pandemic period. However, elevated mortality relative to baseline was also observed in younger age groups during later pandemic phases. Although absolute excess death numbers remained low for individuals younger than 30 years, relative deviations from expected mortality levels were still apparent.

#### 3.2.2 Relative number of Excess Deaths

Although the vast majority of excess deaths occurred in the age group ≥80 years, elevated mortality relative to baseline was also observed in younger age groups. In 2020, excess mortality was largely restricted to the oldest age group, whereas in later years elevated mortality extended progressively into younger age groups. In 2021, a period dominated by the Delta wave, elevated mortality was already apparent in the groups aged 40–79 years. In 2022, elevated mortality relative to baseline was observed in all age groups except children younger than 15 years. Although absolute excess death numbers differed substantially between age groups, relative excess mortality (P-scores) was often within a similar range, typically between 5% and 10%.

Examining the younger groups, the group between 30 and 39 is less affected than the group between 15 and 29. Whereas an absolute number of 280 excess deaths in this group is quite small, a P-score of 7.31% corresponds to a comparatively high relative deviation from baseline expectations. It has to be remarked that other studies support the findings of excess mortality appearing in younger groups as well [23], [21].

For this reason, an additional calculation for this age band was done to get better temporal resolution of mortality effects in this group. The data problem which happens here is that for this age band the deaths are only available in monthly intervals. So, wastewater measurement values had to be aggregated into a monthly interval too and were averaged.

Resulting curves are shown in Figure 4. Baseline is again calculated from the trend 2013-19. Excess is visible for the second half of the year, with peaks of excess deaths in December and September with wastewater peaking in October and December. Thus, this study cannot find a clear temporal alignment between wastewater data and excess deaths in this group.

**Figure 4.**
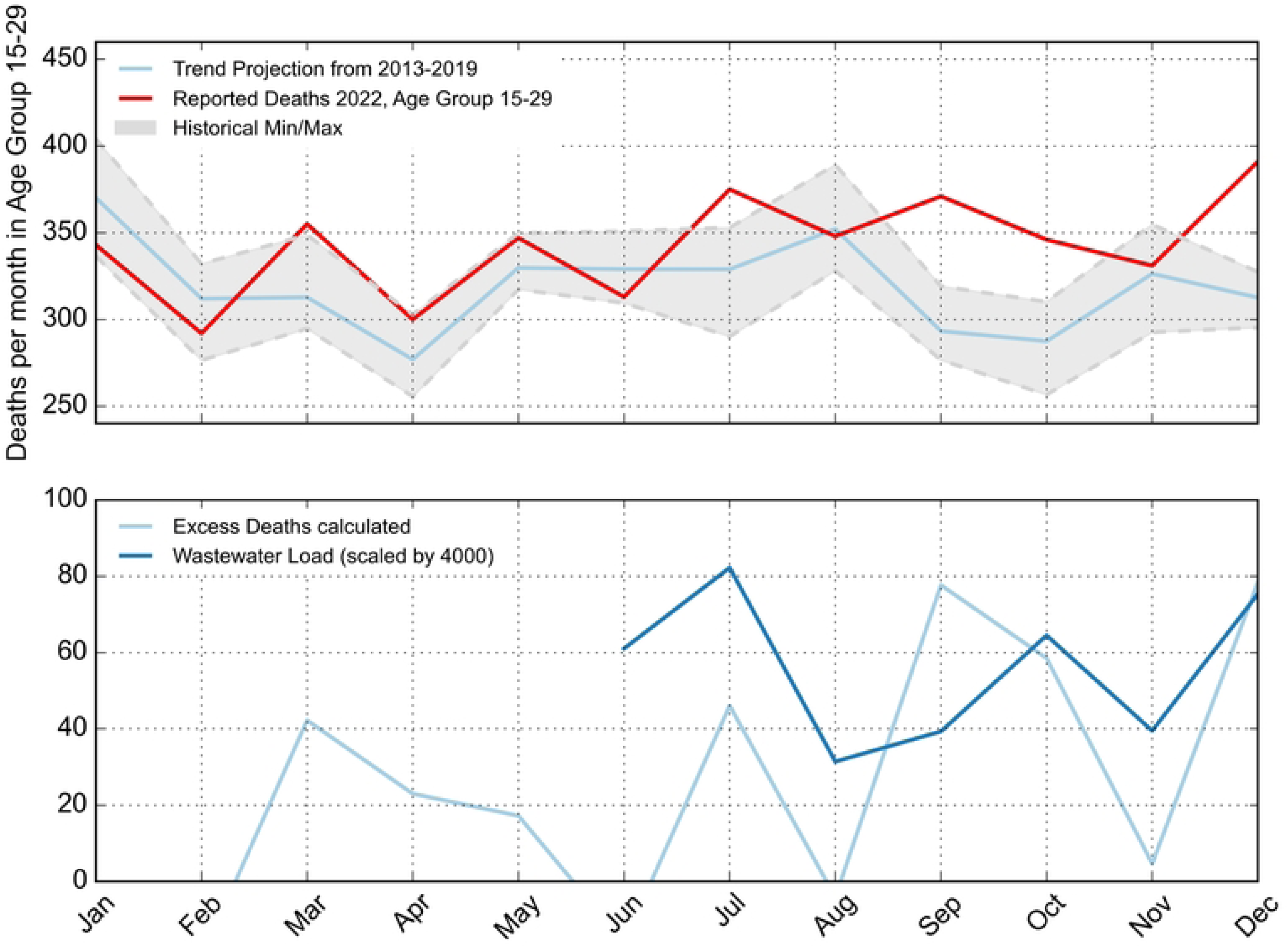
Excess Deaths (monthly values) for the age group 15-29 in the months of 2022. Baseline is calculated from the trend 2013-2019. Reported deaths exceed expected values for September, October and December. Wastewater load is added in monthly averages.

Possible limitations of this straight-forward approach using CDC may be development of the population within demographical changes. Alternative approaches using CMR values show similar results and are intensively discussed in the supplement sections S 9 and S 10. Younger groups show demographic change due to migration. As different approach the trend between 2016 and 2019 may be worth a discussion, which is also done in the supplement. As a result, the P-scores even show stronger effects of excess mortality.

#### 3.2.3 Overall Effect and Age structure

##### 3.2.3.1 Age dependency of Excess Deaths

For each year and mentioned group the absolute number of excess deaths is calculated and normalised to the population in each year, yielding “Risk for Excess Deaths” *R_x_*which is equivalent to the ratio of excess deaths to the population.

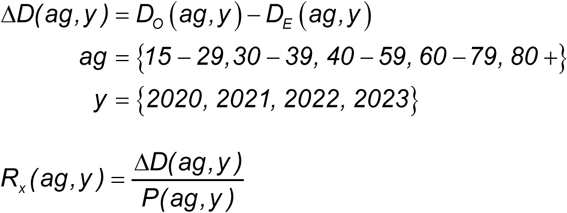

These values are aggregated for the 4 years:

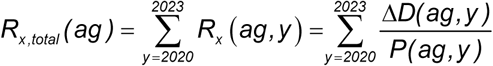

And plotted against the median value of each age bracket, choosing 22.5, 35, 50 and 70 for the three younger groups and 90 for the group older than 80.

The result is shown with a semilogarithmic scale. The logarithmic values of the three older age groups (40-59, 60-79 and 80+) are fitted by a linear dependency. The slope of the logarithmic value can be expressed as:

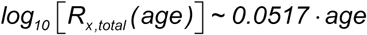

Figure 5 shows this in the upper panel. The lower panel shows the dependency of the registered COVID-19 deaths from the age, as described in the next section.

**Figure 5.**
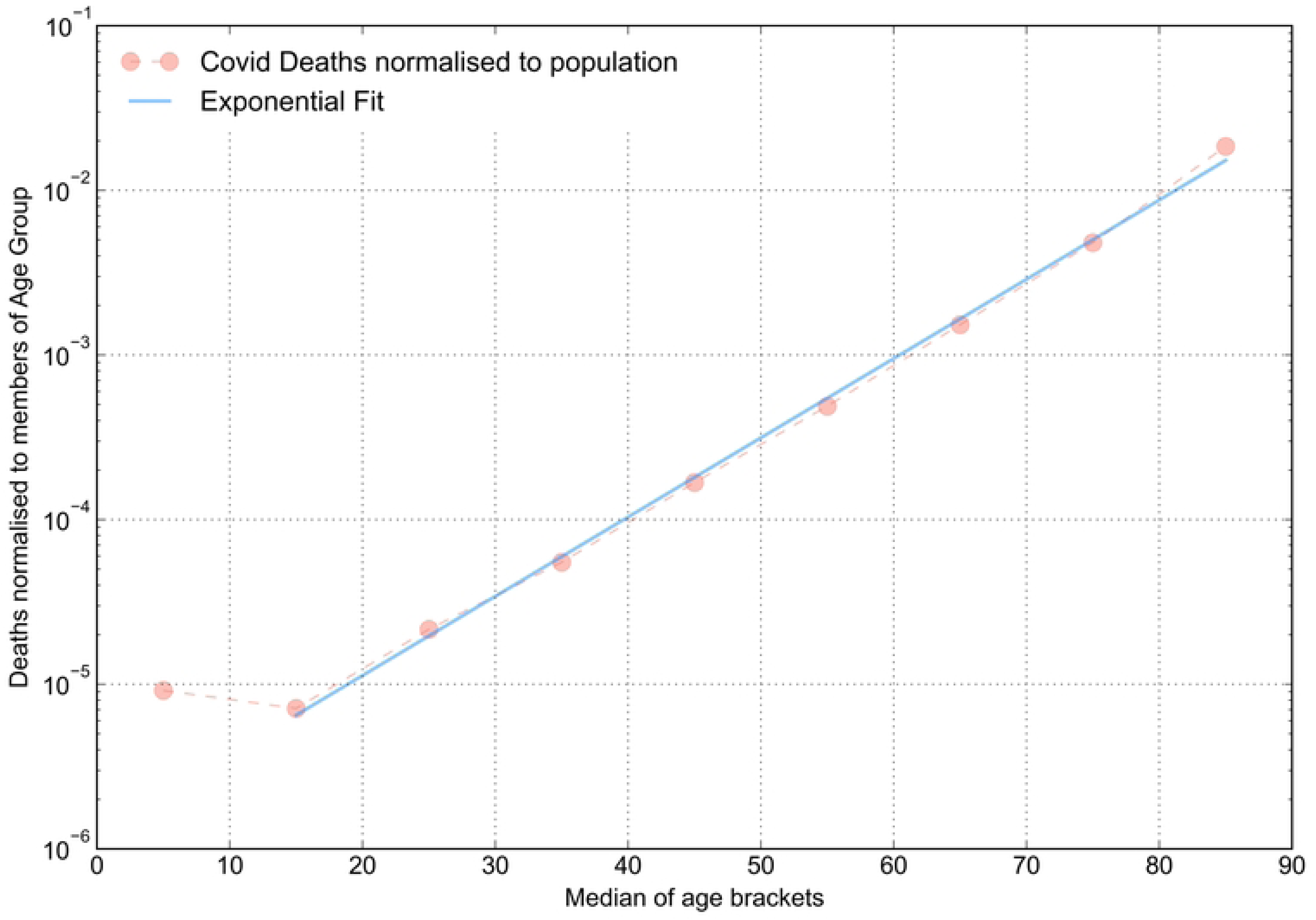
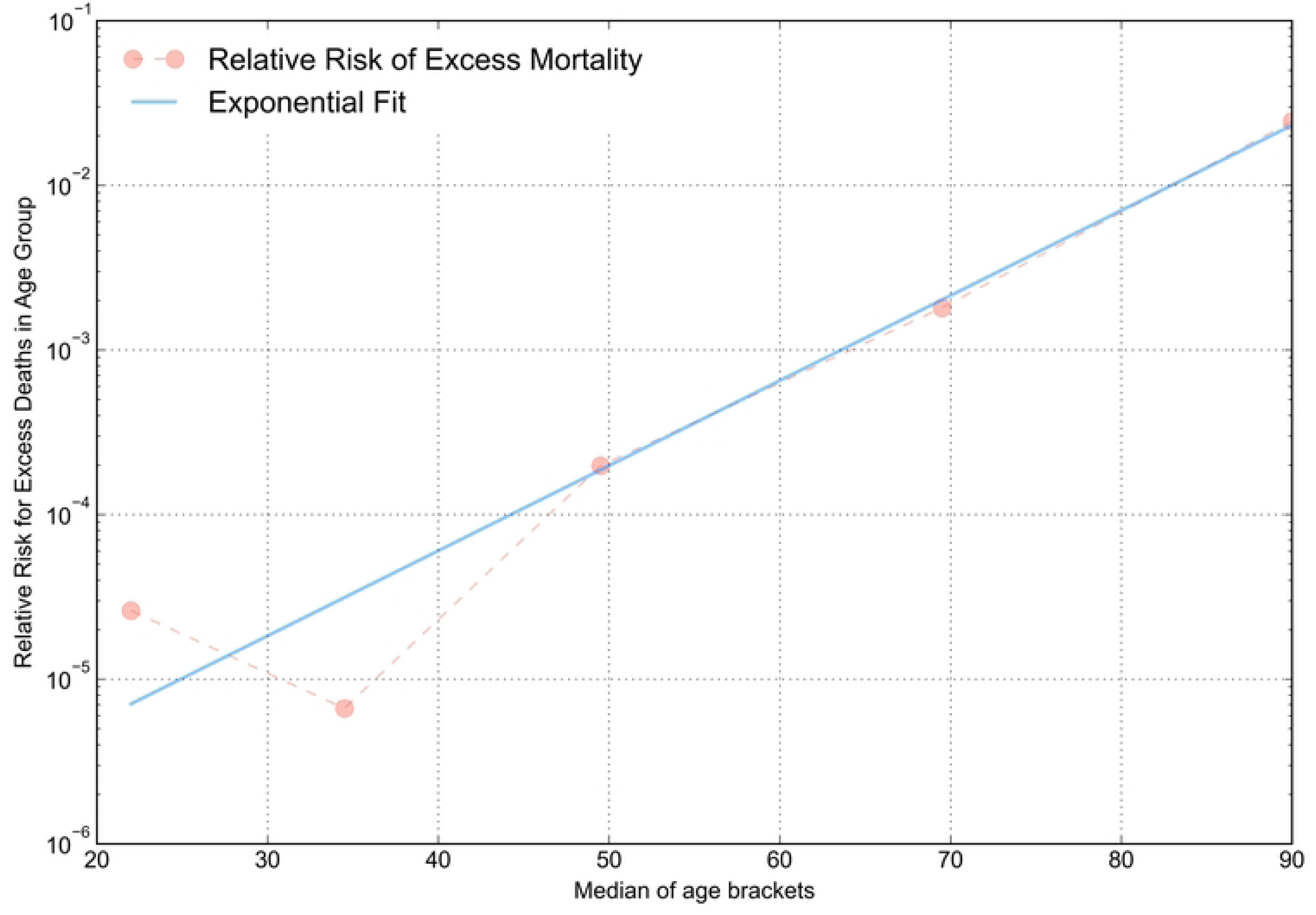
Upper panel: COVID-19 Deaths in age groups registered by RKI, normalised to population size of age groups. Lower panel: „Relative Risk” of excess deaths in age groups (equivalent to excess deaths per population), plotted on a logarithmic y-scale. All values were calculated from the trend 2013-2019. The blue line is a fit of the log10-values of the three groups 40-59, 60-79 and >80. For both quantities, log10-values were fitted linear.

##### 3.2.3.2 Registered COVID-19 Deaths

Age dependent numbers of COVID-19-Deaths, aggregated from the beginning of 2020 until June 2023 were collected from here [31]. I calculated the deaths in 10-year age-brackets normalised to the population:

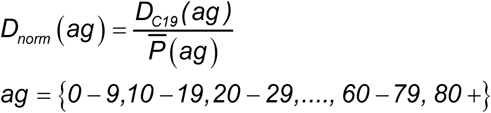

Population number from 2022 [43] was used as approximation for the average over the 4 pandemic years.

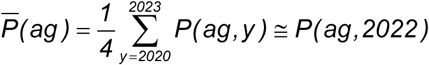

The resulting values are plotted with the y-axis using a log10-scale, fitting the logarithmic values of the inner 7 groups from 15 to 89 years results in surprisingly good fit with a remarkably high regression coefficient of 0.9997. The slope can be expressed as:

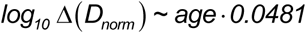

which is in a very good agreement with research results calculating IFR in pre-vaccination era from seroprevalence studies [44], [45].

## 4 Summary and Discussion

This study assesses the effects of excess mortality in Germany between 2020 and 2023. The main findings can be summarized as follows:

Excess mortality in the pandemic years is rising from 2020 to 2022, where it reached its highest level. In 2023, excess mortality declined, indicating a possible return to levels before the emergence of SARS-CoV-2. CDC for 2024 aligns with the pre-pandemic trend used here.

From 2020 to 2022 elevated mortality became increasingly apparent in younger age groups over time. Elevated mortality relative to baseline was observed in 2022 for all age groups except children younger than 15 years. These periods were temporally aligned with waves of reported COVID-19 deaths.

The ratio of all-cause-excess deaths to reported COVID-19 deaths increased for the later years during the Delta– and Omicron-Periods.

COVID-19-mortality has a very clear exponential age dependency which is very consistent with the dependency of IFR from age. In contrast, excess mortality did not show the same monotonic age dependency, with relative excess mortality being higher in the 15–29 age group than in the 30–39 group.

The findings of this study show that periods of excess mortality increasingly exceeded the number of officially reported COVID-19 deaths during later pandemic waves. However, a central question remains: What factors may underlie the observed divergence between excess mortality and officially reported COVID-19 deaths during later pandemic waves? While retrospective analyses such as this one may provide valuable insights on a population level, they also underscore the need for further multidisciplinary research to fully elucidate the underlying causes, particularly concerning younger individuals who experienced increased mortality in 2022. Detailed studies examining their health status prior to death and potential COVID-19 infections in the weeks preceding their deaths could provide critical insights. However, as time passes, the feasibility of answering these questions diminishes, despite their significant implications for both scientific understanding and public health policy.

This study has several limitations. The analysis is based on aggregated population-level mortality data and therefore does not permit causal inference at the individual level. Furthermore, the observed excess mortality can only partially be attributed to specific causes of death. In particular, the largest peak of excess mortality at the end of 2022 coincided with a strong influenza wave, [46] making it impossible to disentangle the relative contributions of SARS-CoV-2, influenza, and other potential factors from the mortality data alone. Estimates of excess mortality also depend on baseline selection and demographic assumptions, particularly for younger age groups with smaller absolute death numbers. In addition, differences between publicly available COVID-19 mortality datasets may contribute to uncertainty in absolute estimates.

## 5 Declaration of ethics, funding and personal interests

This publication uses only publicly available data, so there was no need to consult an ethic commission. I declare that I did not receive any funding for this work and that I have no concurring personal interests. Clinical trial number: not applicable

## 6 Data and Code Availability

All data used in this study originate from publicly available sources, including Destatis, Eurostat, and the Robert Koch Institute (RKI). Detailed references and download links are provided in the manuscript and Supplementary Material.

The scripts used for data processing, statistical analysis, and figure generation are available at: https://github.com/MartinSauter/DE-Excess-Mortality

Archived COVID-19 mortality datasets no longer hosted on the RKI website were accessed via the Internet Archive Wayback Machine, as referenced in the manuscript.

## Data Availability

https://github.com/MartinSauter/DE-Excess-Mortality

## 7 Abbreviations

IFR: Infection Fatality Rate
CDC: Crude Death Count
CMR: Crude Mortality Rate
ASMR: Age-standardised Mortality Rate
NSO: National Statistics Office
destatis: Deutsches Statistisches Bundesamt
RKI: Robert-Koch-Institut
HMD: Human Mortality Database
WMD: World Mortality Dataset
NHS: National Health Service

