## Supplemental Materials for "Excess mortality in Germany during 2020–2023: A descriptive age-stratified analysis"

*Martin Sauter*
*Universität der Bundeswehr München*
*Department of Electrical Engineering and Applied Computer Science*
*Werner-Heisenberg-Weg 39, D-85577 Neubiberg*

#### S 1 Choosing the best regression framework

CDC of Germany since 2009 are shown in Figure S 1, showing a stable rising trend. Demographic systems are changing slowly and so linearization is an appropriate idea for modelling.

Different linear regressions are going back from 2019, beginning in 2016 and back to 2009. The timespan between 2015 and 2019 is an outlier, giving lower predictions for the baseline and higher estimates for excess deaths. Median value for prediction comes from 2013-2019 which will serve as baseline for all other analyses.

The analytical expression for this trend is:

$$D_e(y) \cong 849.880 + 11.328 \cdot (y - 2010)$$

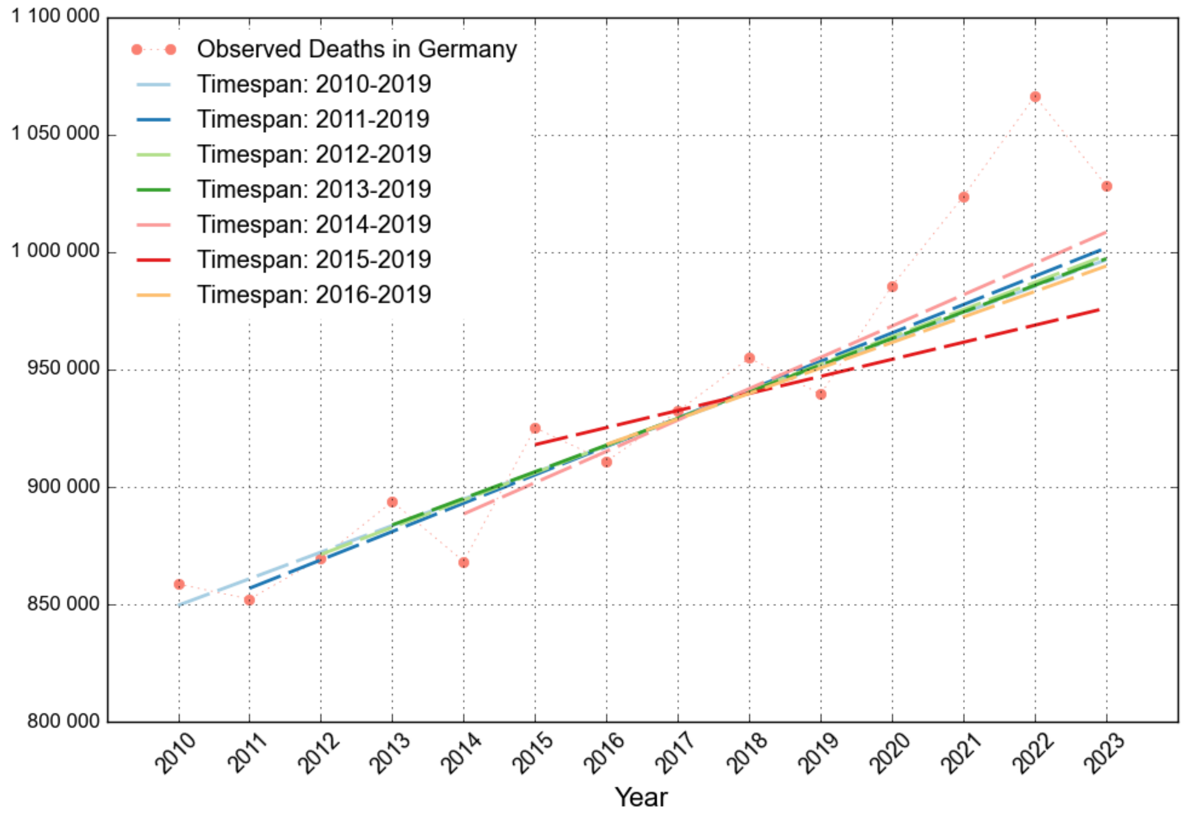


*Figure S 1: Observed Deaths in Germany from 2010 until 2023. Linear Regression is performed with 7 different timespans, each one going back from 2019. Trend 2013-2019 is the most balanced one and will serve as baseline for further discussions.*

The following table summarizes the values from linear regression using different time intervals. Trend 2013-2019 represents the median value for prediction of 2023 and for the slope of the resulting linear function. It also comes closest to the average of all calculated slopes.

*Table S 1: Parameters of linear regression for various time spans in the pre-pandemic years. Second column refers to the number of expected deaths in 2023. This value is the most important one for defining best trend. Red numbers mark median values of slope an intercept resp. the closest ones found, blue numbers the averages found and closest.*

| Timespan | Expected 2023 | M (Slope) | B (Offset) | Min./Max. relative deviation during Timespan (historical Min./Max.) |
|---|---|---|---|---|
| 2010-2019 | 996 696 | 11 310 | 849 669 | -2.97% / +2.09% |
| 2011-2019 | 1 001 751 | 12 068 | 844 866 | -2.77% / +2.21% |
| 2012-2019 | 998 625 | 11 575 | 848 156 | -2.92% / +2.12% |
| **2013-2019** | 997 148 | 11 328 | 849 880 | -3.00% / +2.06% |
| 2014-2019 | 1 008 436 | 13 320 | 835 272 | -2.27% / +2.59% |
| 2015-2019 | 976 121 | 7 261 | 881 725 | -1.56% / +1.60% |
| 2016-2019 | 994 043 | 10 846 | 853 050 | -1.17% / +1.60% |
| Median | 997 148 | 11 328 | 849 669 | - |
| Average | 996 117 | 11 101 | 851 803 | - |

Poisson regression is used in mortality surveillance systems such as EuroMOMO for modelling annual trends [1]. A comparison between linear and Poisson regression is shown in Figure S 2 and Table S 2 Expected deaths for the year 2023 derived from linear regression (LR) and Poisson regression (PR) using different pre-pandemic reference periods ending in 2019. The last column shows the relative difference between both approaches. For all investigated timespans, deviations remained below 0.5%, indicating that the choice between linear and Poisson regression has only a negligible effect on baseline estimates for German mortality data. Both approaches yielded nearly identical baseline estimates, with differences below 0.5% for all investigated reference periods. Because annual mortality increased only gradually during the pre-pandemic period, the exponential growth assumed by Poisson regression exhibits only minimal curvature and closely approximates a linear trend. Consequently, the choice between linear and Poisson regression has only a negligible effect on the resulting excess mortality estimates.

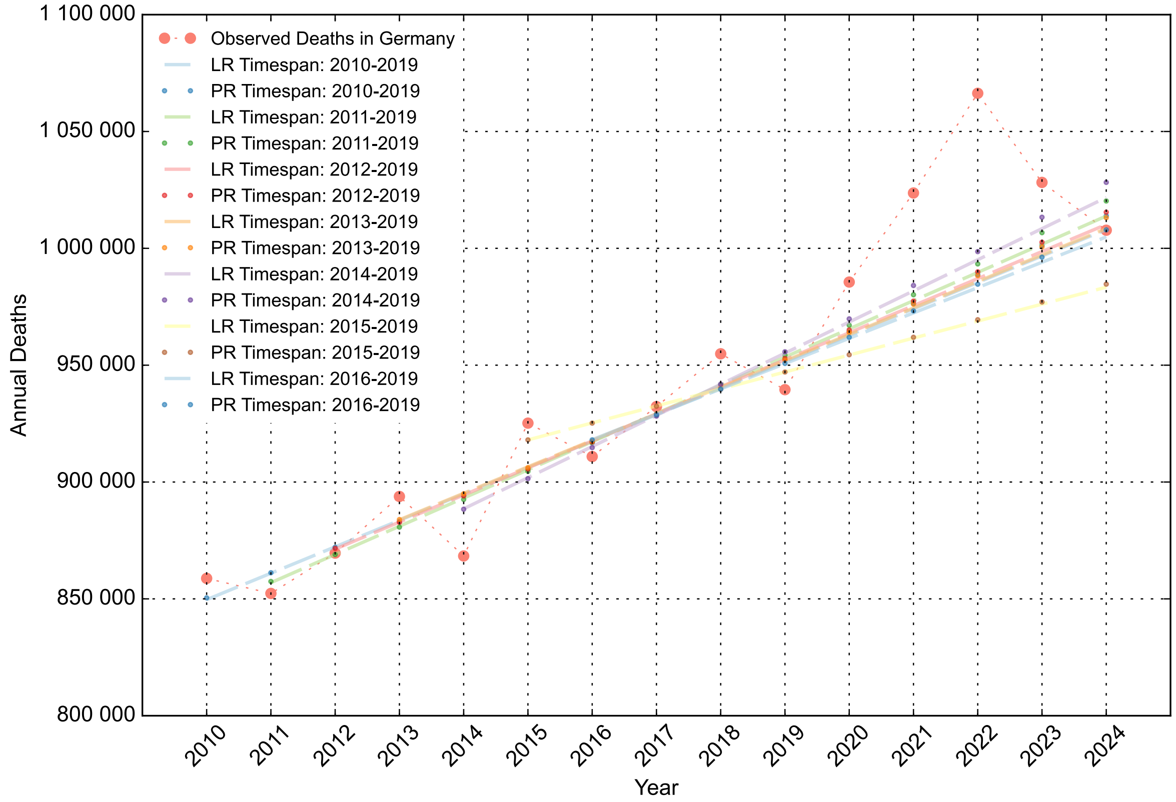


*Figure S 2 Comparison of baseline estimates obtained from linear regression (LR) and Poisson regression (PR) using different pre-pandemic reference periods ending in 2019. Poisson regression is used in mortality surveillance systems such as EuroMOMO. For all investigated timespans, both approaches produced nearly identical projections of expected deaths, with differences below 0.5% for the year 2023 (Table S 2). The close agreement reflects the approximately linear increase in annual mortality during the pre-pandemic period, resulting in only minimal curvature of the exponential trend implied by the Poisson model.*

*Table S 2 Expected deaths for the year 2023 derived from linear regression (LR) and Poisson regression (PR) using different pre-pandemic reference periods ending in 2019. The last column shows the relative difference between both approaches. For all investigated timespans, deviations remained below 0.5%, indicating that the choice between linear and Poisson regression has only a negligible effect on baseline estimates for German mortality data.*

| Timespan | Linear Regression expected 2023 | Poisson Regression expected 2023 | Relative difference between Poisson and linear regression |
|---|---|---|---|
| 2010-2019 | 996 696 | 1 001 263 | 0.46% |
| 2011-2019 | 1 0017 51 | 1 006 705 | 0.49% |
| 2012-2019 | 998 625 | 1 002 617 | 0.40% |
| **2013-2019** | 997 148 | 1 000 622 | 0.35% |
| 2014-2019 | 1 008 436 | 1 013 306 | 0.48% |
| 2015-2019 | 976 121 | 977 034 | 0.09% |
| 2016-2019 | 994 043 | 996 220 | 0.22% |

Models based on actuarial approaches using life-tables were also evaluated and compared with the linear baseline used in this study. In particular, the model of De Nicola and Kauermann [2] projected a markedly steeper increase in expected deaths than any of the intervals used for linear regression, especially steeper than the result coming from linear regression between 2016 and 2019.

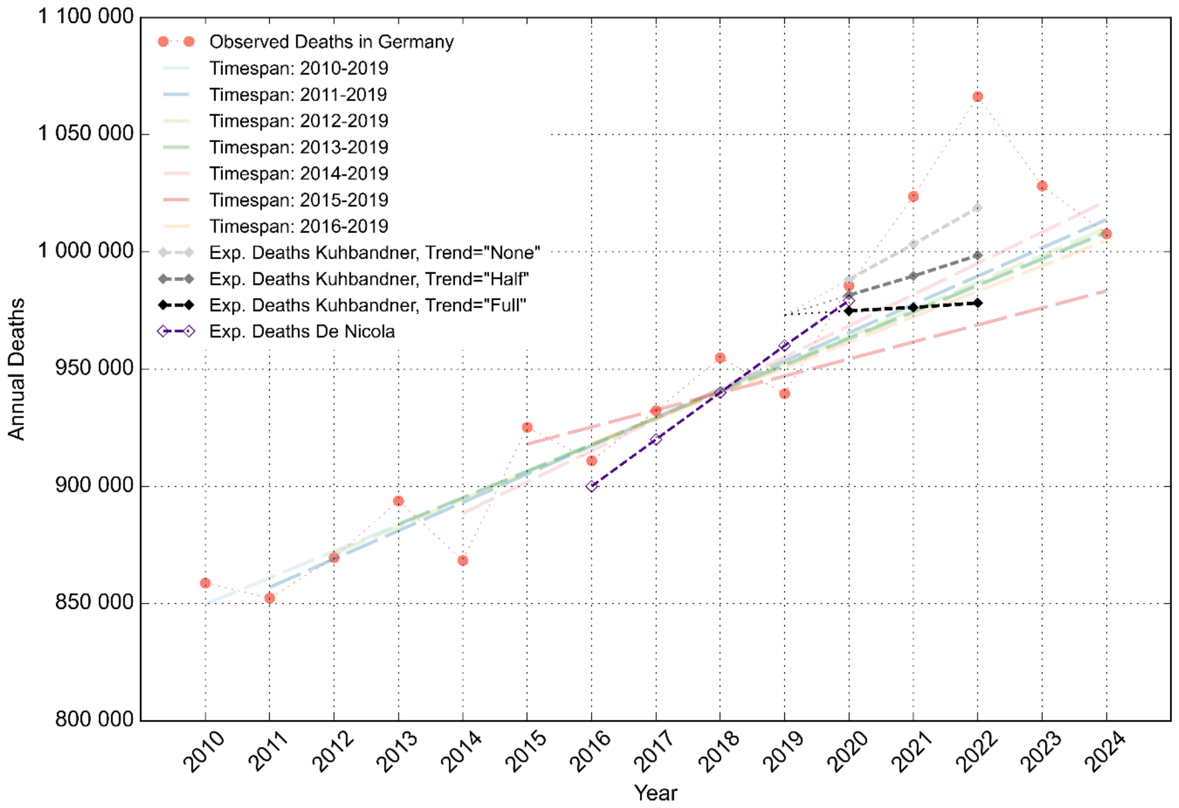


*Figure S 3 Comparison of expected annual deaths derived from alternative baseline models. Linear regressions fitted to different pre-pandemic reference periods are shown together with the actuarial approaches proposed by Kuhbandner and Reitzner and by De Nicola and Kauermann. While the linear models form a comparatively narrow range of projections, the actuarial models produce substantially different extrapolations beyond 2019. Observed annual deaths in Germany are shown for reference.*

The approach of Kuhbandner and Reitzner [3] employs three alternative baseline trends derived from the actuarial life table DAV 2004 R (Deutsche Aktuarvereinigung mortality table for annuity insurance) [4]. When extrapolating these three trends backwards to 2019, all three trends meet at reference values in 2019 of approximately 973 000 expected deaths. However this value exceeds the observed mortality in 2019 (939 000 deaths) by approximately 3.6%, corresponding to a difference of roughly 34 000 deaths. Consequently, the resulting baseline estimates are already elevated at the end of the calibration period, before any extrapolation into the pandemic years is performed.

Furthermore, three alternative trend specifications ("none", "half", and "full") are proposed, but the rationale for selecting one specific trend for excess mortality estimation is not clearly defined. As a result, estimated excess mortality remains dependent on the chosen baseline specification.

Although linear regression represents a comparatively simple modelling approach, it produced baseline estimates that remained stable across different reference periods, showed close agreement with Poisson regression, and were broadly consistent with subsequently observed mortality levels. Taken together, these findings support its use as a transparent and robust baseline model for the present analysis.

#### S 2 Deviations from previous years vs. prediction intervals

The very simple method to use maximum and minimum deviations for previous years as indicator if deaths are declared as excess or not is here compared with prediction intervals coming from the standard linear model. Here, the prediction boundary is 68%, which represents the one-sigma boundary value of a normal distribution. The area within this PI is shaded in light grey. Both limits com quite close together. The chosen method gives asymmetrical intervals for upper and lower bounds, whereas PIs are symmetrical. This is caused by the underlying assumption of a normal distribution of the data, which may not be the case within here.

When using a 68% PI, the years of 2020 and 2023 do not show to have a "significant" number of excessed deaths.

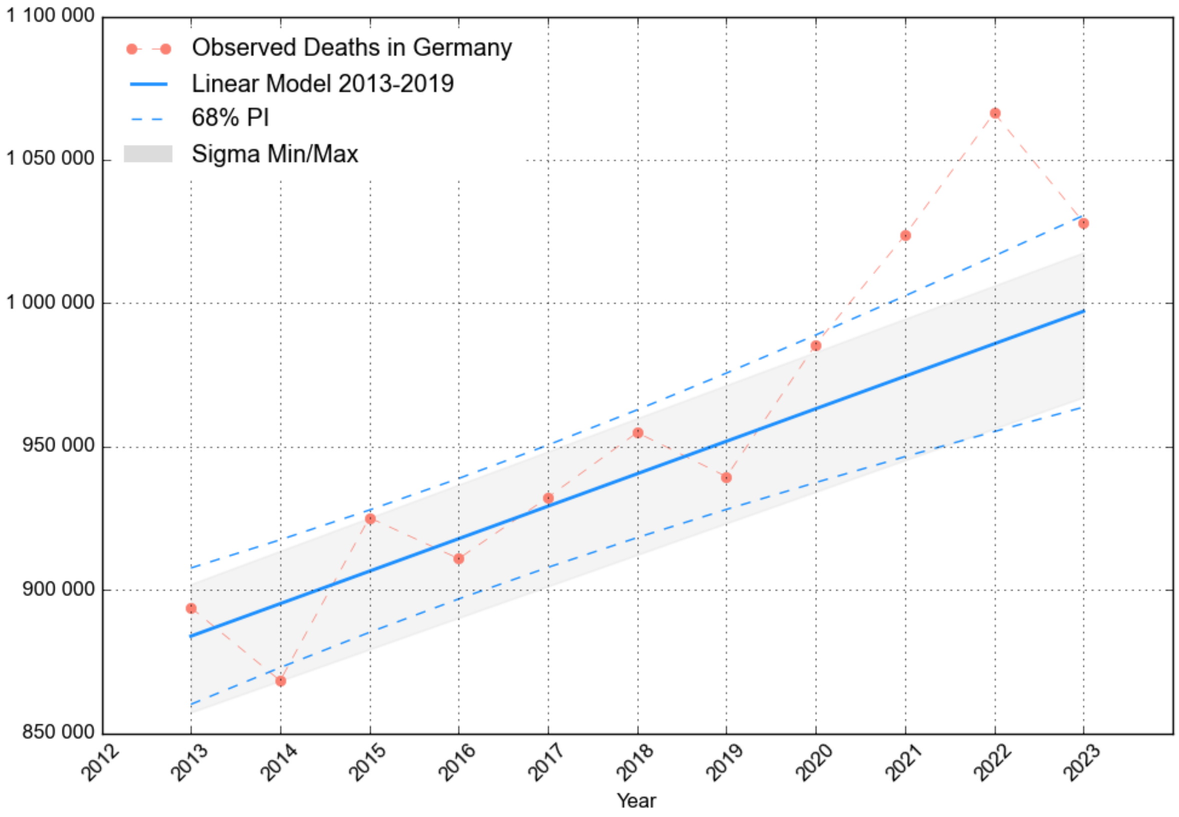


*Figure S 4: Grey shaded area shows the chosen minimum/maximum estimation resulting from the previous years vs. Prediction intervals arising from linear models. Prediction intervals plotted here represent ±1σ in a normal distribution.*

#### S 3 Decomposition into subgroups

In general terms, a linear regression applies to a set of a dependent variable $y_i$ and an independent variable $x_i$ with the index $i$ running from 1 to N. Slope $m$ and intercept $b$ in the regression $y=m_y \cdot x + b_y$ are calculated from the variables as follows (here I denote by the subindex $y$ that these values belong to the variable set $(x_i,y_i)$ ):

$$m_y = \frac{\sum_{i=1}^{N}(x_i - \overline{x})(y_i - \overline{y})}{\sum_{i=1}^{N}(x_i - \overline{x})^2} \qquad b_y = \overline{y} - m \cdot \overline{x}$$

With $\overline{x}$ and $\overline{y}$ as average of the sets. If we assume that the dependent variable $y_i$ can always be split into two groups (e.g. observed deaths of all people partitioned into deaths of men and women) we write for all valid indices $i$:

$y_i = u_i + v_i$

If we apply linear regression now to the subsets $u$ and $v$ we get similar formulas like above, using the letters $u$ and $v$ as result for the two subsets:

$$m_u = \frac{\sum_{i=1}^{N}(x_i - \bar{x})(u_i - \bar{u})}{\sum_{i=1}^{N}(x_i - \bar{x})^2} \qquad b_u = \bar{u} - m_u \cdot \bar{x}$$

$$m_v = \frac{\sum_{i=1}^{N}(x_i - \bar{x})(v_i - \bar{v})}{\sum_{i=1}^{N}(x_i - \bar{x})^2} \qquad b_v = \bar{v} - m_v \cdot \bar{x}$$

If we look at the sum of the two fits and add them, we will see that their sum is identical to the fit of $y_i$=$u_i$+$v_i$ :

$$\begin{aligned} u + v &= m_u x + b_u + m_v x + b_v \\ &= (m_u + m_v)x + (b_u + b_v) \end{aligned}$$

$$\begin{aligned} m_u + m_v &= \frac{\sum_{i=1}^{N}(x_i - \bar{x})(u_i - \bar{u})}{\sum_{i=1}^{N}(x_i - \bar{x})^2} + \frac{\sum_{i=1}^{N}(x_i - \bar{x})(v_i - \bar{v})}{\sum_{i=1}^{N}(x_i - \bar{x})^2} \\ &= \frac{\sum_{i=1}^{N}(x_i - \bar{x})\left[(u_i - \bar{u}) + (v_i - \bar{v})\right]}{\sum_{i=1}^{N}(x_i - \bar{x})^2} \\ &= \frac{\sum_{i=1}^{N}(x_i - \bar{x})\left[\left(\overbrace{u_i + v_i}^{y_i} - \left(\overbrace{\bar{u} + \bar{v}}^{\bar{y}}\right)\right)\right]}{\sum_{i=1}^{N}(x_i - \bar{x})^2} \\ &= m_y \end{aligned}$$

$$\begin{aligned} b_u + b_v &= \bar{u} - m_u \cdot \bar{x} + \bar{v} - m_v \cdot \bar{x} \\ &= \bar{u} + \bar{v} - (m_u + m_v) \cdot \bar{x} \\ &= \bar{y} - m_y \cdot \bar{x} \end{aligned}$$

And we can conclude that the sum of the two regressions for each subgroup *u* and *v* is identical to the regression of *y=u+v*, so that values based from predictive extrapolations will also sum up.

#### S 4 Effect of different baselines on time resolution

Figure S 5 compares weekly baseline estimates for 2020 derived from the reference periods 2013–2019 and 2015–2019. The two models produce highly similar results throughout most of the year. The largest deviations occur during calendar weeks 1–10, whereas differences during the summer heat period (CW 32–36) are comparatively small. The difference within calendar weeks 1 to 10 is 6461 reported deaths, accounting for more than 70% of the total difference between the corresponding annual baseline estimates (8826 deaths). This indicates that most of the discrepancy between the two models originates from differences in mortality during the first weeks of the year.

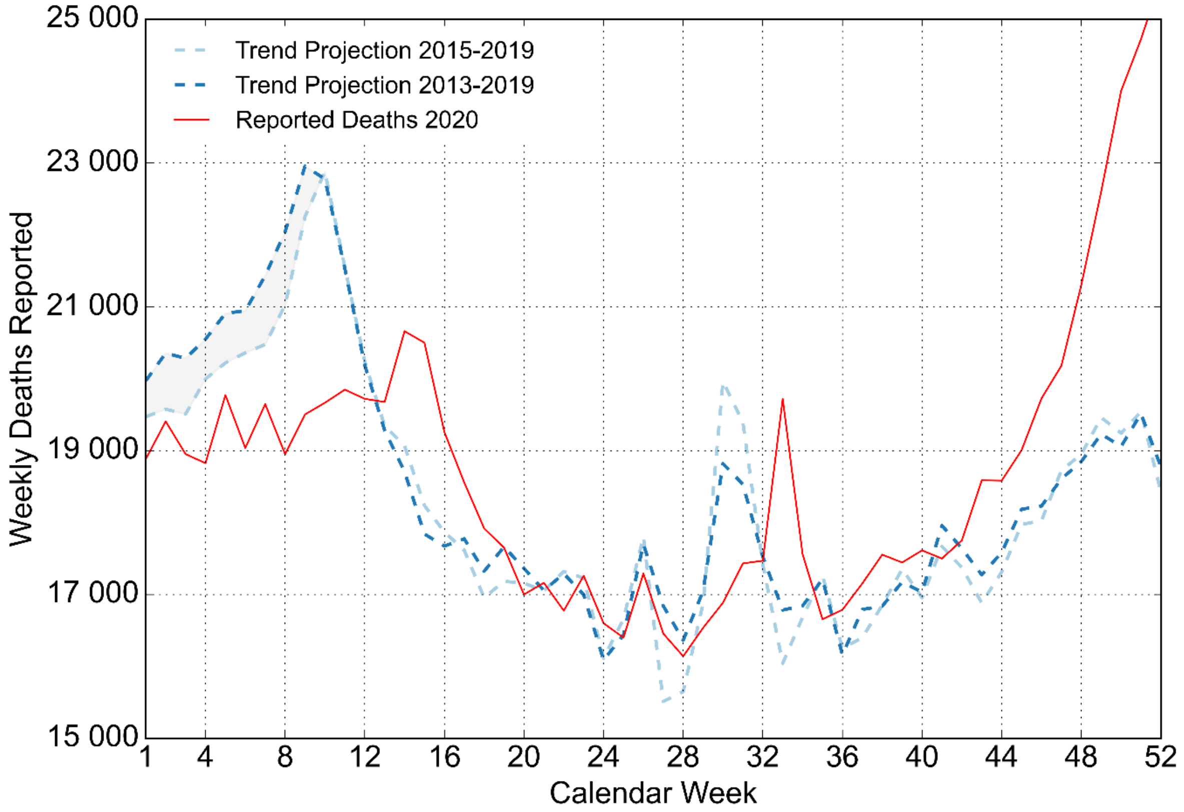


*Figure S 5: Comparison of weekly mortality baselines for 2020 derived from the periods 2013–2019 and 2015–2019. Observed deaths in 2020 are shown in red. The main discrepancy between the two baseline models occurs during calendar weeks 1–10 (grey shaded area), corresponding to 6461 deaths. Most of the difference between the annual projections can therefore be attributed to differences in influenza mortality contained in the respective reference periods.*

#### S 5 The problem of Data sources for COVID-19 fatalities

A data analysis reveals quite a difficult situation regarding data of certified COVID-19 fatalities. I list here the source and the type of data they showed.

Until mid of 2023 RKI presented two different types of excel sheets for the public, containing confirmed fatalities. Both of these are not hosted anymore on RKI's website but can still be accessed via the wayback machine. The first one [5] gives fatalities aggregated over years in different age groups, this one is used for calculating the age structure of the COVID-19 fatalities. Temporal resolution is here only available weekly.

The second one [6] gives weekly and monthly resolution but is not ideal for age structure, because for small numbers (in the younger groups) below four there is no precise number given, but instead only the string "<4" which gives some uncertainties.

The third one was introduced in 2023 when a GitHub archive with machine readable .csv-files was created. Three different files show deaths in age groups, federal states and for whole Germany [7]

But when trying to assess the total over the years, these three files are inconsistent, especially the third one gives a much lower number for 2020. As the date here is labelled as "Date of report", this is possibly not corrected for reporting delay.

The fourth source comes from Destatis and originates from the death cause statistics [8]. The dataset includes the two rows "TDU -18, COVID-19 virus detected" and "TDU-19, COVID-19, virus not detected" and were summarized together for an overview.

| Source | 2020 | 2021 | 2022 | 2023 |
|---|---|---|---|---|
| RKI “Klinische Aspekte” (until 06/23) [5] | 52 747 | 65 966 | 47 908 | incomplete |
| RKI “COVID-19_Todesfälle” (until 06-23) [6] | 41 835 | 73 352 | 48 701 | incomplete |
| RKI github archive (agg. from daily data) [7] | 33 071 | 78 854 | 49 540 | 18 644 |
| Destatis [8] | 39 758 | 71 331 | 52 357 | 25 768 |

*Table S 3: Compilation of different sources for certified COVID-19 deaths*

### S 6 ASMR calculations

For various scenarios, the age-standardised mortality rates (ASMR) were calculated as well. I used here:

- ASMR with 5-year age bands, using the German population in 2020 (DE2020) as standard population
- ASMR with 10-year age bands using the standard populations WHO2015, ESP2013 and DE2020

Again, trend is calculated from 2013 to 2019, resulting in a falling line, which represents rising life expectation. The resulting P-scores show that the first calculation matches with the P-scores calculated from Crude Death Count (CDC) quite good. Using 10-year age bands generally reduces the resulting P-scores, particularly for the year 2020.

The observed sensitivity arises because excess mortality in 2020 was concentrated almost entirely in the oldest age groups, as shown in section 3.2 of the main paper. Standard populations with a younger age structure, such as WHO2015, assign lower weights to these groups and therefore yield substantially lower excess mortality estimates than standard populations that more closely resemble the age structure of Germany in 2020.

| Standard Population | P-score 2020 | P-score 2021 | P-score 2022 | P-score 2023 | Historical Min./Max. |
|---|---|---|---|---|---|
| Values from CDC | 2.33% | 5.05% | 8.17% | 3.11% | -3.00% / 2.06% |
| DE2020, 5-year bands | 2.33% | 4.92% | 8.26% | 3.80% | -2.68% / 1.83% |
| DE2020, 10-year bands | 1.09% | 3.39% | 6.78% | 3.25% | -2.62% / 2.34% |
| ESP2013, 10-year bands | 0.97% | 3.44% | 6.57% | 3.00% | -2.60% / 2.19% |
| WHO2015, 10-year bands | 0.68% | 3.57% | 6.23% | 2.66% | -2.57% / 2.00% |

*Table S 4: P-scores for different calculations of age-standardised mortality rates.*

### S 7 Population Count and Development

The model used here assumes a continuous change (either growth or decline) of the population. I will take a closer look of the population numbers given by German NSO since 2005 here. These numbers are available in the GENESIS Database [9], the following graph shows the development of the German population since 2009 (For each year, the value represents the number reported by German NSO for the of the 31.12. of the previous year)

We can see interesting discontinuities in this graph, which may anyway be explained in a plausible manner.

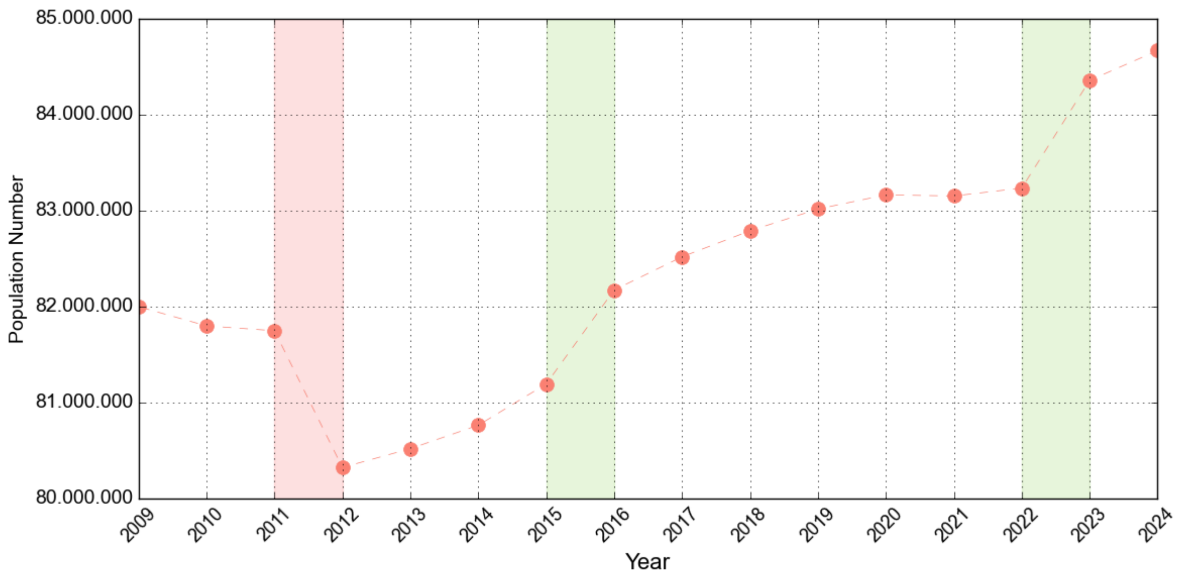


*Figure S 6: Counted Population in Germany. The shaded areas mark sudden inclines and declines. These are explained in the text.*

The first drop from 2011 to 2012, shaded in light red is a correction by a new census performed in 2011 [10]. This means, this is a completely artificial drop coming from the fact that population count is merely based on estimates and is not as precise as counting death numbers.

The second two inclines represent migration effects. In 2015 more than one million Immigrants came to Germany due to middle east refugee crisis, the origin of the immigrants were mainly Syria, Iraq and Afghanistan. The vast majority of this group were young men in the ages between 20 and 30.

The incline from 2022 to 2023 represents another refugee group, mainly younger families coming from Ukraine and fleeing the Russian invasion [11]. This migration effect also affected mainly the younger groups. At the same time, a new census was done which would lead to another correction of population data down by more than one million inhabitants. Anyway, the numbers used here are based on the values assessed by the previous census and prognosis from 2011. It is very difficult to decide how correct these values are. As a simple approach I assume that the numbers from 2011 and 2022 are correct and I calculate a “corrected population count” by a linear interpolation between the values from 2013 and 2022.

Effects of sudden changes of the population should be eliminated by using crude mortality rate (CMR) instead of death numbers. I will also use the similar linear regression method to calculate a relevant baseline based on this metric and compare the resulting p-scores. The following graph shows the data including linear fit and relative deviations from the baseline. Assuming the explained limitation of correctness of inhabitant count, I also calculate a “corrected mortality rate” (CorrMR) from death numbers divided by these values. For both values I do again a linear regression and show resulting p-Scores.

The results are shown as a side-by-side comparison in Table S 5. The results are very interesting: P-scores of CDN match very well with the P-scores of CorrMR-Values. CMR P-scores tend to be even higher for 2022 and 2023. This can be explained by not correcting the resulting drop of CMR for 2022.

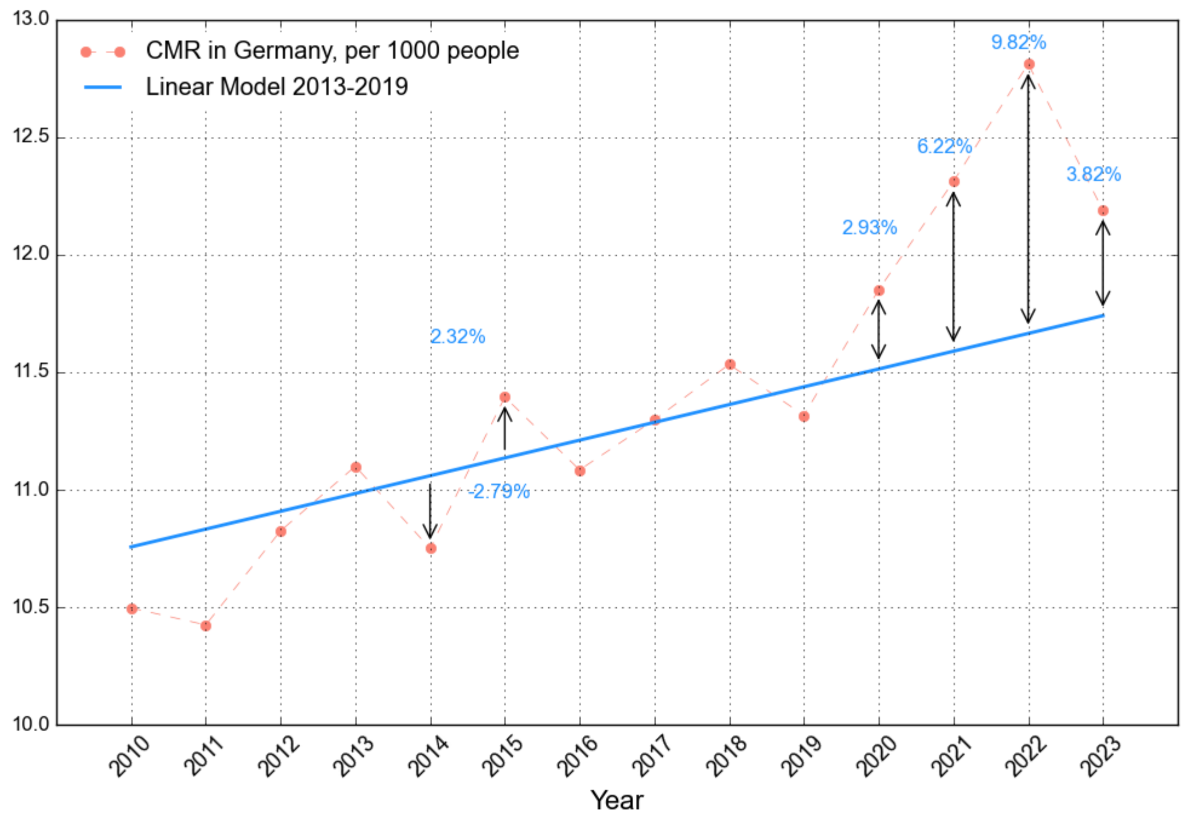


*Figure S 7: Linear Fit for Crude Mortality Rate (CMR) with reference timespan 2013-2019. Calculated P-scores for the years 2020 to 2023 are added in the graph, the values and the pattern are quite similar to calculations from CDC (Figure S 1). Side-by-side comparison of P-scores is shown below in Table S 5.*

All P-scores show the same qualitative result and give a strong support for the basic findings already shown in Section 2 of our Paper.

*Table S 5: Calculated P-scores for Death Numbers (DN), Crude (CMR) and Corrected (CorrMR) Mortality Rates. All regressions were performed from 2013 to 2019. All values show the same outcome in qualitative terms. P-scores of CorrMR and DN match very well, P-scores from CMR are higher than from CDN, the correction puts them in the direction of CDN-P-score values. The red colour is an indication that the deviation from expectation is higher than in the previous years.*

| Calendar Year | P-score of CDC | P-score of CMR | P-score of Corrected MR |
|---|---|---|---|
| **2020** | 2.33% | 2.93% | 2.28% |
| **2021** | 5.05% | 6.22% | 4.97% |
| **2022** | 8.17% | 9.82% | 8.05% |
| **2023** | 3.11% | 3.82% | 2.96% |
| **Min/Max prev. years** | -3.00/+2.06% | -2.09%/+2.32% | -3%/+2.07% |

#### S 8 Excess Mortality in Age Groups

The methodology presented above allows to break down total excess deaths into partial groups. For this purpose, I split the population into age bands, which were (more or less) 20 years wide. Due to the database of the German National Statistics office I chose 6 age bands ranging from 0-15, 25-29, 30-39, 40-59, 60-79 years and older than 80 years. The main intent of this choice to assure a continuous trend in population. I will show both trend of CDN and corresponding population size and discuss inconsistent p-scores resulting from CDN and CMR.

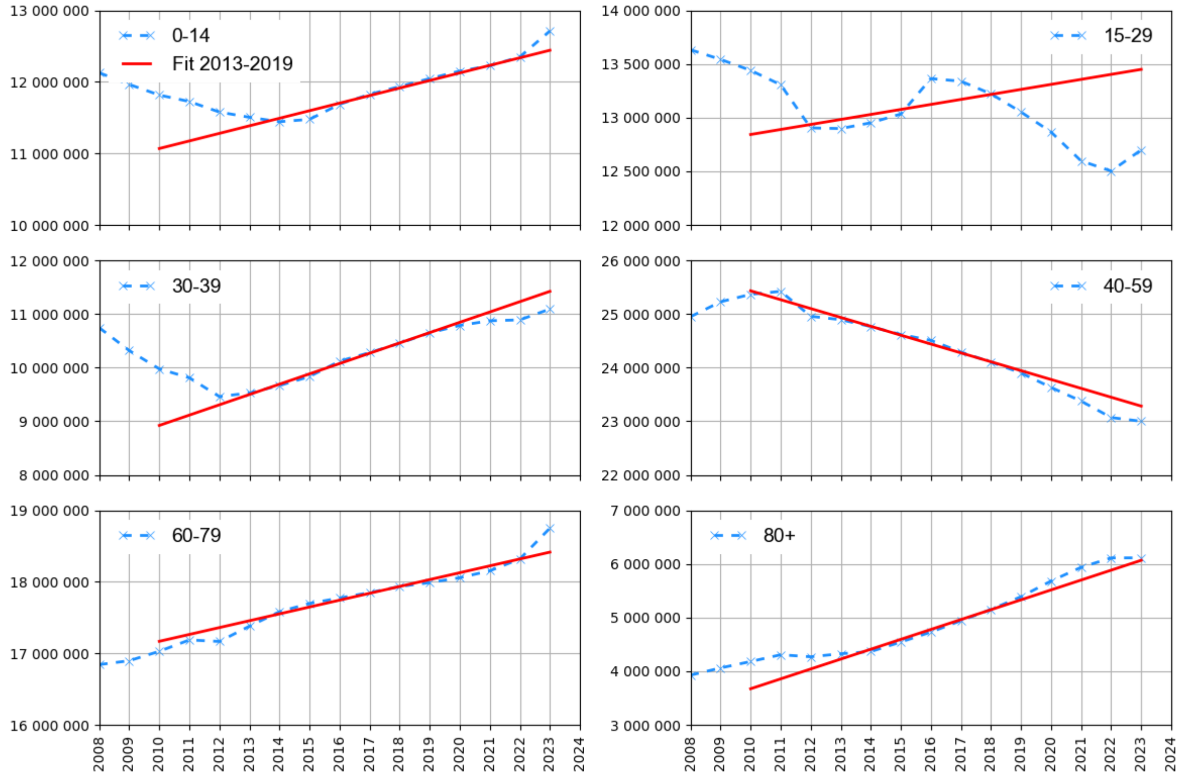


*Figure S 8: Population numbers in the age bands used for the calculations here. The 20-year age bands try to find population branches with a more or less continuous change in the group sizes. Crude mortality rates are also calculated in the following chapters in order to check whether population effects may affect calculated excess mortality.*

#### S 9 CMR and CDC of Age group 80+

A notable discrepancy emerges when comparing excess mortality estimates based on CDC and CMR in the population aged 80 years and older. Given the substantial excess mortality observed in this age group during 2020 and 2021, both measures would be expected to show qualitatively similar patterns. However, while CDC-based estimates indicate pronounced excess mortality, the corresponding CMR-based estimates remain comparatively low. This discrepancy coincides with a strong increase in the estimated size of the population aged 80 years and older between 2020 and 2022, despite the occurrence of considerable excess mortality during the same period. Population figures published by Destatis are based on demographic projections between census years rather than direct annual counts, and therefore remain subject to uncertainty, particularly for specific age groups. To assess whether such uncertainty could plausibly explain the observed discrepancy, an alternative population trajectory was constructed by linear interpolation between the census-based population estimates of 2013 and 2022.

This correction is not intended as an alternative population estimate, but rather as a sensitivity analysis. The resulting “corrected mortality rates” (CorrMR) yield P-scores much closer to those obtained from CDC, indicating that uncertainty in the population estimates of the oldest age group may plausibly account for a substantial part of the discrepancy between CDC- and CMR-based excess mortality estimates.

*Table S 6: P-scores derived from Crude Death Counts (CDC), Crude Mortality Rates (CMR) and Corrected Mortality Rate (CorrMR) for the years from 2020 to 2023 in the group of age older than 80. CorrMR was calculated by using the interpolation from Figure S 9 instead of the numbers provided by destatis. Values from CMR show no rising above the level of previous years, whereas CDC-P-scores do this very well. This obvious contradiction can be resolved by fitting population numbers, which corrects values closer to values calculated from CDC. Numbers in red indicate excess mortality effects.*

| Age Group 80+ | 2020 | 2021 | 2022 | 2023 | Min/Max of previous years |
|---|---|---|---|---|---|

| | | | | | |
|---|---|---|---|---|---|
| **P-score from CDC** | 4.43% | 6.28% | 10.94% | 3.91% | -3.77% / +2.22% |
| **P-score from CMR** | 1.76% | 2.76% | 8.01% | 4.79% | -2.74% / +3.22% |
| **P-score from CorrMR** | 4.94% | 7.14% | 12.27% | 5.61% | -3.80% / +2.30% |

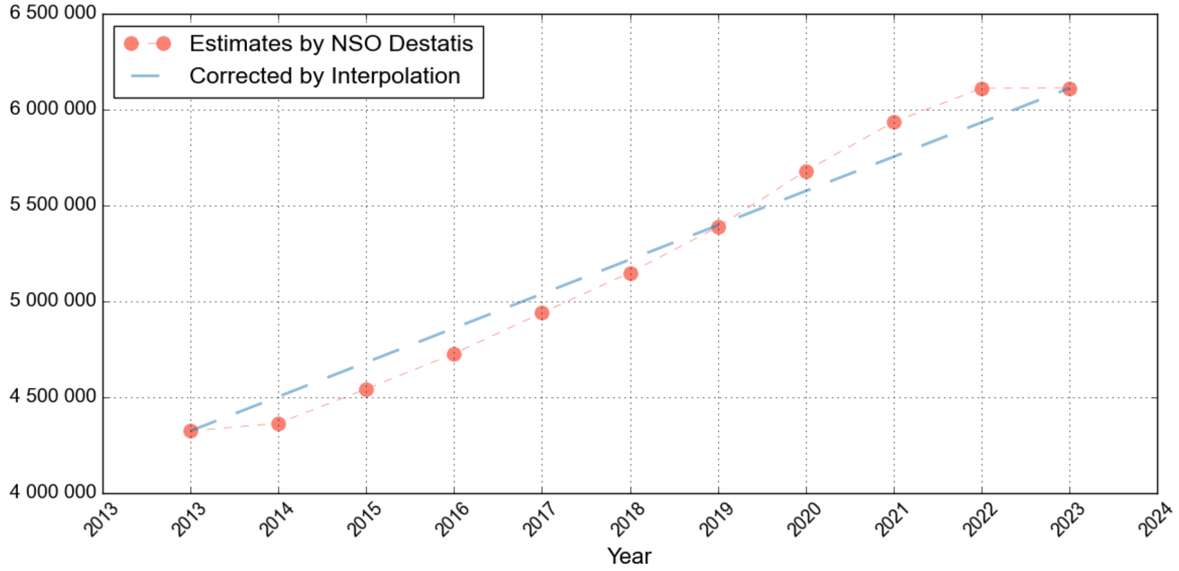


*Figure S 9: Population of Inhabitants of age over 80 given by German NSO destatis. Linear interpolation between 2013 and 2022 is shown as well as blue line.*

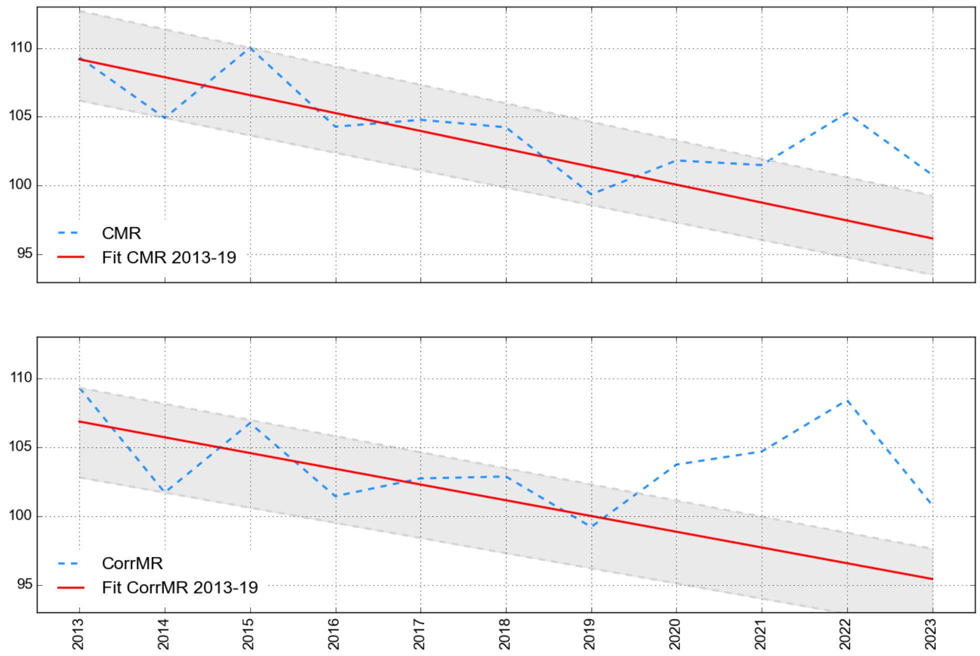


*Figure S 10: Results of linear trends for the age group >80 years using CMR per 1000 (upper panel). The blue line shows calculated CMR, red the trend from the years 2013-19. Using CMR, excess mortality in this group does not occur for 2020 and 2021, which is apparently contradictive to the results from CDC. In the lower panel, the corrected mortality rate (CorrMR) which comes from the assumption that population numbers are strictly linear changing between 2013 and 2022. This assumption leads to relative values of excess mortality similar to these resulting from CDC.*

#### S 10 Comparison CDC with CMR for the middle-age groups

When examining these numbers, we can see a change in the age dependency of excess deaths in the younger groups as well. For 2020 we do not see these groups affected at all. For 2021 and 2022 excess deaths are visible in the groups 60-79 and 40-59. Both markers (CDC and CMR) show the same: P-scores differ slightly, but the effect is clear. When effects differ, the origin is caused by a population number deviating from a previous trend.

*Table S 7: P-scores derived from Crude Death Counts (CDC) and Crude Mortality Rates (CMR) for the years from 2020 to 2023 in the groups between 30 and 79. These groups are affected only in the later years of the pandemic, there are no excess deaths visible for 2020. Group 30-39 is only affected for 2022.*

| Age Group | Marker | 2020 | 2021 | 2022 | 2023 | Min/Max of previous years |
|---|---|---|---|---|---|---|
| **60-79** | **P-score (CDC)** | -0.53% | 3.18% | 4.76% | 2.93% | -2.14% / +1.97% |
| | **P-score (CMR)** | -0.10% | 3.67% | 4.91% | 1.29% | -2.31% / +1.67% |
| **40-59** | **P-score (CDC)** | 0.23% | 5.25% | 2.43% | -1.48% | -1.81% / +1.43% |
| | **P-score (CMR)** | 0.82% | 6.24% | 3.99% | -0.44% | -1.80% / +1.47% |
| **30-39** | **P-score (CDC)** | -0.29% | 0.52% | 2.81% | -2.04% | -3.01% / +2.61% |
| | **P-score (CMR)** | 0.31% | 2.15% | 6.22% | 1.16% | -2.73% / +3.12% |

An exception occurs for the group of 30-39. Here excess deaths happen only in 2022. P-score of CDC in 2022 (2.81%) is only very small above the maximum of the previous years (2.61% in 2015), but P-score of CMR is much higher. Both markers do not show any excess for the other years.

#### S 11 Comparison of different trends for the younger groups

##### *Age group 15-29*

Effects in this group are even more pronounced than in the group 30-39. There is no excess in 2020, but later happening from 2021 to 2023. Checking the population (Figure S 8b.) shows an interception in population trend in 2015, also in CDC a visible increase at this point can be seen. When using the trend from 2013 to 2019 for extrapolation this leads to different numbers than using the trend from stable population measurements 2016 to 2019. The trend 2016-2019 shows a stronger effect in excess deaths than the original one 2013-2019. As discussed in the first section this does not affect the overall outcome, but is important to understand for a proper assessment of excess mortality in the younger groups.

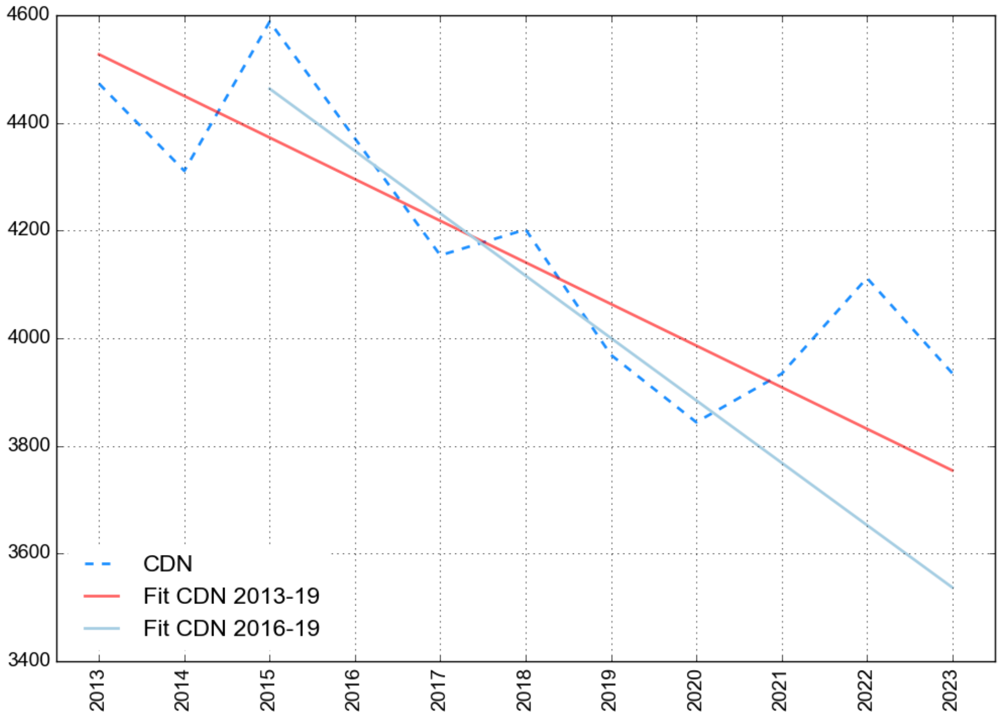


*Figure S 11: CDC in the age group 15-29 in the relevant timespan 2013 to 2023. The linear fits (light blue and red) refer to different periods used for the linear extrapolation. Population growth due to migration is shown in Figure S 8 and points out to the fact that the trend from 2016 to 2019 is more reliable than the original one (2013-2019). Using the shorter timespan leads to higher values of calculated relative excess deaths, a comparison is listed below in Table S 8 .*

*Table S 8: P-scores for Age Group 15-29, here calculated from the original trend (2013-2019) and the shorter trend 2016-2019.*

| Age Group 15-29 | 2020 | 2021 | 2022 | 2023 | Min/Max of previous years |
|---|---|---|---|---|---|
| **P-score from CDC (Trend 16-19)** | -1.03% | 4.41% | 12.61% | 11.30% | -1.843% / +2.08% |
| **P-score from CDC (Trend 13-19)** | -3.57% | 0.64% | 7.32% | 4.81% | -3.14% / +4.93% |

##### *Children between 0-14*

Evaluating this group faces the same questions as the previous group. When applying a trend from 2013 to 2019 there is no excess mortality visible at all, extrapolation leads instead to high deficit numbers for all pandemic years.

When applying a trend from 2016 to 2019, the overall picture changes. For the first year (2020) an obvious mortality deficit occurs, for the further years CDC are above expectation level. For 2022 and 2023, a rise of CDC above expectation level can be seen. But the conclusion here is that there is no clearly visible excess mortality in this group, even not in a qualitative point of view.

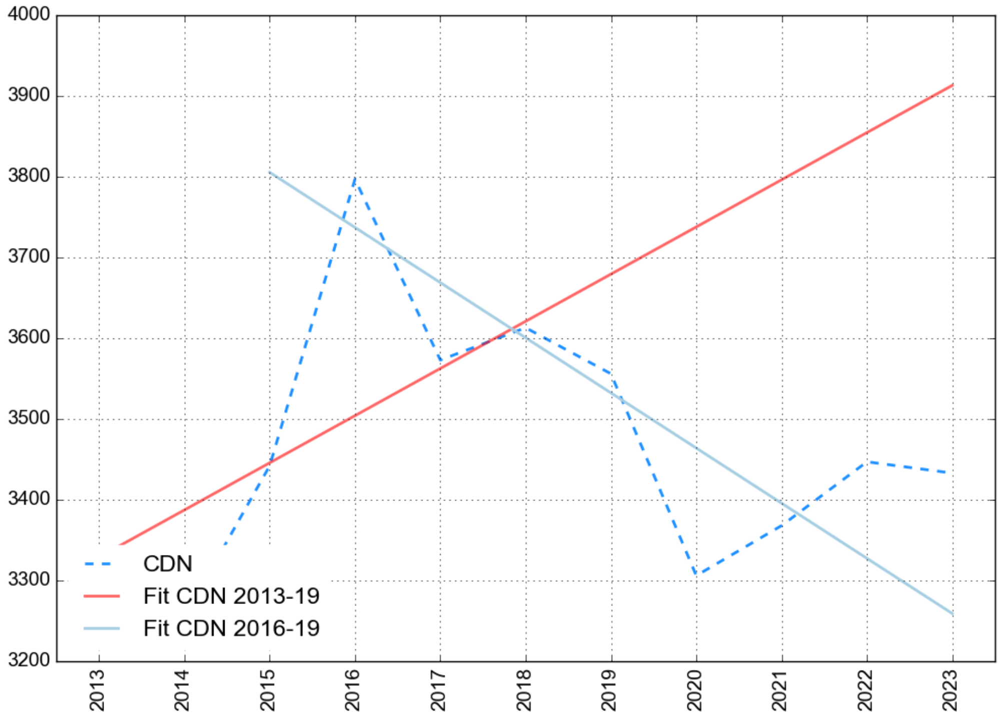


*Figure S 12: CDC in the age group 0-14 in the relevant timespan 2013 to 2023. The linear fits (light blue and red) refer to different periods used for the linear extrapolation. Population growth due is stable in the years before 2022. Different time intervals lead to completely different P-scores. A comparison is listed in Table S 9.*

*Table S 9: P-scores for Age Group 0-14, here calculated from Trends from 2016 to 2019.*

| Age Group 15-29 | 2020 | 2021 | 2022 | 2023 | Min/Max of previous years |
|---|---|---|---|---|---|
| **P-score from CDC (Trend 2013-2019)** | -11.55% | -11.28% | -10.58% | -12.27% | -3.91% / +8.36% |
| **P-score from CDC (Trend 2016-2019)** | -4.56% | -0.82% | 3.59% | 5.34% | -2.61% / +1.60% |

Bibliography


1. EuroMOMO Methods. EuroMOMO; Available: https://www.euromomo.eu/how-it-works/methods

2. De Nicola G, Kauermann G, Höhle M. On assessing excess mortality in Germany during the COVID-19 pandemic. AStA Wirtsch- Sozialstatistisches Arch. 2022;16: 5–20. doi:10.1007/s11943-021-00297-w

3. Kuhbandner C, Reitzner M. Estimation of Excess Mortality in Germany During 2020-2022. Cureus. 2023 [cited 23 Sept 2024]. doi:10.7759/cureus.39371

4. Deutsche Aktuarvereinigung. Herleitung der DAV-Sterbetafel 2004 R für Rentenversicherungen. Köln: Deutsche Aktuarvereinigung; 2023. Available: https://aktuar.de/content/PDF/Fachwissen/2023-06-28_DAV-Richtlinie_Herleitung_DAV2004R.pdf

5. RKI. Todesfälle aus Klinische Aspekte (via web.archive.org). 9 July 2023. Available: https://web.archive.org/web/20230709201850/https://www.rki.de/DE/Content/InfAZ/N/Neuartiges_Coronavirus/Daten/Klinische_Aspekte.xlsx?__blob=publicationFile

6. Robert Koch-Institut. COVID-19 Todesfaelle (RKI, via web.archive.org). Available: https://web.archive.org/web/20230714161929/https://www.rki.de/DE/Content/InfAZ/N/Neuartiges_Coronavirus/Projekte_RKI/COVID-19_Todesfaelle.xlsx?__blob=publicationFile

7. RKI. COVID-19-Todesfälle in Deutschland. doi:10.5281/zenodo.15030264

8. Destatis. Germany, years, causes of death, sex. Available: https://www-genesis.destatis.de/datenbank/online/statistic/23211/table/23211-0001

9. destatis. Population: Germany, reference date, age. Available: https://www-genesis.destatis.de/datenbank/online/statistic/12411/table/12411-0005

10. Zensus 2022. In: Zensus 2022: 82,7 Millionen Einwohnerinnen und Einwohner [Internet]. Available: https://www.zensus2022.de/DE/Aktuelles/PM_Zensus_2022_Bevoelkerungszahl_Ergebnisveroeffentlichung.html

11. Rat für Integration e.V. 2022 Geflüchtete Statistik. In: Asylzahlen 2022 [Internet]. Available: https://mediendienst-integration.de/artikel/die-wichtigsten-asylzahlen-2022.html
